# A systematic review and meta-analysis of glyphosate-based herbicide exposure and risk of non-Hodgkin’s lymphoma

**DOI:** 10.64898/2026.02.26.26347184

**Authors:** Joel J. Gagnier, John O’Connor

## Abstract

**Background:** Glyphosate-based herbicides are among the most widely used agricultural chemicals globally. Concerns regarding their carcinogenic potential, particularly in relation to non-Hodgkin’s lymphoma (NHL), persist despite multiple prior systematic reviews and meta-analyses. However, these reviews have important methodological limitations and inconsistent analytic decisions, limiting confidence in their conclusions.

**Objective:** To conduct a systematic review and meta-analysis of studies examining the association between glyphosate-based herbicide exposure and risk of NHL and its subtypes, while addressing methodological and analytic shortcomings of prior syntheses.

**Methods:** We searched MEDLINE and EMBASE (to February 26, 2026), supplemented by reference list review. Eligible studies included cohort, case-control, and pooled analyses and risk of bias was assessed using recommended criteria. Random- and fixed-effects meta-analyses were conducted using inverse-variance methods. Heterogeneity was evaluated using Cochran’s Q and I² statistics. Publication bias was assessed using standard and contour-enhanced funnel plots. Sensitivity analyses addressed overlapping cohorts, hazard ratio inclusion, exposure definitions, and model overfitting (events-per-variable considerations). Certainty of evidence was graded using GRADE.

**Results:** Seventeen publications were identified, representing 20 unique study populations; after accounting for overlap, 10 primary datasets were included in quantitative synthesis. Five studies were assessed as low risk of bias, four as moderate risk, and one as high risk. For ever exposure, the random-effects model across yielded an odds ratio (OR) of 1.11 (95% CI: 0.98–1.27; I²≈53%). In sensitivity analyses ORs ranged from 1.19 to 1.23. For the highest exposure categories, the random-effects model demonstrated a statistically significant association (OR≈1.38; 95% CI: 1.00–1.90; I²≈61%). Sensitivity analyses strengthened the association (OR≈1.47; 95% CI: 1.04–2.06). Analyses incorporating alternative cumulative exposure metrics yielded similar associations (OR≈1.33–1.45). Publication bias assessments revealed evidence of small-study effects in some models, though contour-enhanced analyses suggested that not all asymmetry was attributable to selective publication. Certainty of evidence was moderate for highest exposure analyses and low-to-moderate for ever-exposure analyses.

**Conclusions:** Associations with ever exposure to glyphosate are modest and sensitive to analytic decisions and higher levels of exposure are consistently associated with increased odds of NHL. Findings are robust across multiple sensitivity analyses addressing overlapping data, exposure classification, and model overfitting.

## Introduction

Glyphosate is an organophosphonate herbicide that is widely used [1]. Individuals that use glyphosate-based herbicides may be heavily exposed during application, mostly by dermal and inhalation exposures [2]. Given the ubiquitous nature of glyphosate in our environment and many foods, there have been concerted efforts to examine its safety. In particular, the risk of non-Hodgkin’s lymphoma has been examined across multiple epidemiologic studies [e.g.4-11]. Furthermore, there have been several systematic reviews on the topic [3,11,13,19,20,27] most having important flaws and all being out of date, not incorporating the most recent evidence on the topic.

In table 1, we provide a detailed assessment of quality of the reviews using the AMSTAR-2 criteria [48]. All reviews were judged to be critically low meaning that they had more than one critical flaw making their risk of bias high. Therefore, the results of all existing systematic reviews and meta-analyses are potentially biased and their results are therefore not trustworthy. Furthermore, the most recent of these systematic reviews was done over five years ago and therefore do not include more recent publications [e.g.,47,57,58].

**Table 1:** Assessment of systematic reviews using AMSTAR-2 Criteria.

| ASMTAR-2 Criteria | Study |  |  |  |  |  |
| --- | --- | --- | --- | --- | --- | --- |
|  | Schinasi, 2014 | Chang & Delzell, 2016 | Zhang, 2019 | Donato, 2020 | Kabat, 2021 | Boffetta, 2021 |
| 1. Did the research questions and inclusion criteria for the review include the components of PICO? | Y | Y | Y | Y | N | Y |
| 2. Did the report of the review contain an explicit statement that the review methods were established prior to the conduct of the review and did the report justify any significant deviations from the protocol? | N | N | N | Y | N | N |
| 3. Did the review authors explain their selection of the study designs for inclusion in the review? | Y | Y | Y | Y | Y | Y |
| 4. Did the review authors use a | N | PY | N | Y | N | Y |
| comprehensive literature search strategy? |  |  |  |  |  |  |
| 5. Did the review authors perform study selection in duplicate? | N | Y | N | Y | N | Y |
| 6. Did the review authors perform data extraction in duplicate? | N | N | N | N | N | N |
| 7. Did the review authors provide a list of excluded studies and justify the exclusions? | Y | N | Y | N | N | N |
| 8. Did the review authors describe the included studies in adequate detail? | Y | Y | Y | Y | N | Y |
| 9. Did the review authors use a satisfactory technique for assessing the risk of bias (RoB) in individual studies that were included in the review? | N | PY | Y | N | N | N |
| 10. Did the review authors report on the sources of funding for the studies included in the review? | N | Y | N | N | N | N |
| 11. If meta-analysis was performed, did the review authors use appropriate methods for statistical combination of results? | Y | Y | Y | N | Y | N |
| 12. If meta-analysis was performed, did the review authors assess the potential impact of RoB in individual studies on the results of the meta-analysis or other evidence synthesis? | N | N | N | N | N | N |
| 13. Did the review authors account for RoB in primary studies when interpreting/discussing the results of the review? | N | Y | N | N | N | N |
| 14. Did the review authors provide a satisfactory explanation for, and discussion of, any heterogeneity observed in the results of the review? | Y | Y | Y | Y | Y | Y |
| 15. If they performed quantitative synthesis did the review authors carry out an adequate investigation of publication bias (small study bias) and discuss its likely impact on the results of the review? | N | Y | Y | Y | N | Y |
| 16. Did the review authors report any potential sources of conflict of interest, including any funding they received for conducting the review? | Y | Y | Y | Y | Y | Y |
| Overall Confidence in the results of the review | Critically Low | Critically Low | Critically Low | Critically Low | Critically Low | Critically Low |

In addition to these flaws, the meta-analyses frequently chose to include effect estimates from the individual studies that were adjusted for a large number of variables, frequently choosing the effect estimate (e.g., OR) that was “the most” adjusted, meaning including the largest number of variables. But, the authors of these reviews did not consider the lack of power and therefore the high risk of spurious (chance) findings in those regression models which can be considered over-fitted models. Overfitted regression models result in untrustworthy effect estimates [e.g., 50-53]; there is much empirical evidence of how overfitting causes false findings. Therefore, in addition to the other methodologic flaws in these papers they also had analytic flaws that result in the findings being invalid and therefore not trustworthy. Given the drawbacks in all meta-analyses to date, the fact that recent papers have been published that are not included in these prior meta-analyses, there was a strong and convincing rationale for carrying out an up-to-date rigorous systematic review and meta-analysis of the literature in this area.

## Methods

### Protocol registration

We prepared a protocol prior to completing this review but it was not registered in PROSPERO due to the existence of several other similar reviews underway. Our a-prior, time-stamped protocol is available upon request.

### Inclusion criteria

Observational study design (cohort, case-control, cross-sectional, population-based studies, pooled analyses), includes estimates of risk, or data to calculate those estimates, for NHL and any type of exposure to glyphosate-based herbicides, participants of any age, studies in any language.

### Search methods

We performed a literature search in Medline (PubMed version) from 1970 to February 26, 2026, reviewed reference lists from relevant reviews and from included studies. The search terms used were as follows: PubMed example search terms: (glyphosat* OR pesticide [MeSH] or herbicides [MeSH]) AND (lymphoma, non-Hodgkin [MeSH] OR lymphoma [tiab] OR non–Hodgkin [tiab] OR non–hodgkins [tiab] OR lymphoma[tiab] OR lymphomas[tiab] OR NHL OR cancer OR cancers) AND (“occupational exposure”[MeSH] OR occupational exposure [tiab] OR occupational exposures[tiab] OR farmers [MeSH] OR farmer OR applicators OR applicator OR agricultural workers OR agricultural worker or workers or worker). We also performed a literature search in EMBASE from its inception February 26, 2026 using the following search strategy: (’glyphosate’/exp OR glyphosate OR ’pesticide’/exp OR pesticide OR ’herbicides’/exp OR herbicides) AND (’lymphoma, non-hodgkin’/exp OR ’lymphoma, non-hodgkin’ OR ((’lymphoma,’/exp OR lymphoma,) AND ’non hodgkin’) OR non–hodgkin OR non–hodgkins OR ’lymphoma’/exp OR lymphoma OR ’lymphomas’/exp OR lymphomas OR nhl OR ’cancer’/exp OR cancer OR ’cancers’/exp OR cancers) AND (’occupational exposure’/exp OR ’occupational exposure’ OR ’occupational exposur’ OR ((’occupational’/exp OR occupational) AND exposur) OR ’occupational exposures’ OR ((’occupational’/exp OR occupational) AND exposures) OR ’farmers’/exp OR farmers OR ’farmer’/exp OR farmer OR ’applicators’/exp OR applicators OR ’applicator’/exp OR applicator OR ’agricultural workers’/exp OR ’agricultural workers’ OR (agricultural AND workers) OR ’agricultural worker’/exp OR ’agricultural worker’ OR (agricultural AND (’worker’/exp OR worker)) OR workers OR ’worker’/exp OR worker) AND (cohort OR ’case control’). We also searched reference lists from isolated articles and relevant review articles.

### Data collection and extraction

One individual performed inclusion and exclusion with a random sample of citations cross-checked separately and independently for inclusion by a second individual. All included studies were abstracted, reviewed and summarized.

### Study risk of bias assessment

The risk of bias (ROB) of the included studies (case-control and cohort studies) was evaluated using the using the Newcastle Ottawa Scale (NOS) [27]. For pooled analyses of epidemiologic studies there are no known risk of bias tools or accepted guidance, therefore we used other related recommendations to guide assessment of these studies [e.g., 48,49]. We used a cutoff of 70% or greater of the items fulfilled as low risk of bias, of between 40% and 69% as moderately high risk of bias, and of between 0 and 39% as very high risk of bias. These categories were predetermined and were used in a follow-up subgroup analyses and total study ROB score was used in meta-regression analyses to explore the influence of study quality on meta-analytic risk ratios. All risk of bias assessments for all included studies were done in duplicate, separately and independently, by two trained individuals and disagreements were resolved by consensus.

### Measures of treatment effect

Study results were analyzed using primarily STATA/MP 14.2. Effect estimates for dichotomous outcome events were expressed as ratios of risks, rates or odds and/or differences of risk or rates, depending on the type of analysis.

### Statistical methods

Summary estimates of the intervention effect were combined across studies of similar interventions and outcomes, using standard methods of meta-analysis [29], provided the data and reported findings are suitable for that purpose. Separate meta-analyses were used for different exposure classifications and outcomes where possible. Meta-analyses, fixed and random effects models, where appropriate, were used to obtain an overall estimate of the intervention effect using inverse-variance weighting.

### Exploration of heterogeneity

Heterogeneity was assessed using three methods: 1) graphical displays of study-specific and overall effect estimates (forest plots); 2) The Cochran’s chi-squared test for the presence of statistical heterogeneity, e.g., where p < 0.10 indicates significant heterogeneity); 3) the I^2^ statistic, where I^2^ > 0.50 may indicate substantial heterogeneity [30]. Meta-regression analysis was used to predict study-specific effect estimates as a function of study-specific indicators of population characteristics (e.g., age, sex, exposure length, exposure intensity / route, concomitant exposures, and comorbidities), study design, and methodologic features (ROB scores) as noted above where possible. The goal is to account for variation in the estimated effect across studies. When the amount of residual heterogeneity not explained by the predictors is judged to be small, fixed-effects models were fitted to the summary data, which assume the true effect, conditional on covariates, is the same in all studies. When the amount of residual heterogeneity was substantial, unexplained variation across studies were treated as random effects in the model. To assess the robustness of our results, sensitivity analyses were done for varying reported effect estimates from the individual studies or overlapping studies, as described below.

### Evaluation of publication bias

Funnel plots, both standard and contour-enhanced, were used to detect signs of publication bias, the phenomenon whereby positive findings are more likely than negative findings to be published [31] and other small study effects [32].

### Grading of the evidence

The GRADE criteria were used to provide an overall grading of the quality of the evidence for each outcome across studies [33]. A GRADE table was also produced. The GRADE assessment was performed separately and independently by two trained individuals.

### Sensitivity analyses

Several meta-analyses include the data from Leon 2019 as well as Poh 2022, which only reported hazards ratios, which may be considered equivalent to ORs, though not exact, since there were a low number of events and the HR was close to 1.0. To test the impact of this decision, these studies were excluded in sensitivity analyses to determine the robustness of results. As well varying analyses were done due to overlapping samples in several papers.

Specifically, the Leon paper include the AHS data up to December 31, 2011 for Iowa and December 31^st^ 2010 for North Carolina, whereas, the Andreotti paper included AHS data up to 2013 for Iowa and December 31^st^ 2012 for North Carolina and was the most up to date data from AHS. Therefore, a sensitivity analyses removing Leon and Poh but including Andreotti was therefore completed.

## Results

### Literature search

From the Pubmed search we found 1,131 citations and from the EMBASE search 1,500 citations. After reviewing reference lists, deleting duplicates and applying the inclusion and exclusion criteria this resulted in 17 unique papers, though with several being updated analyses and overlapping cohorts associated with prior papers. There were no disagreements on the inclusion of papers between the two reviewers.

### PRISMA flow Diagram

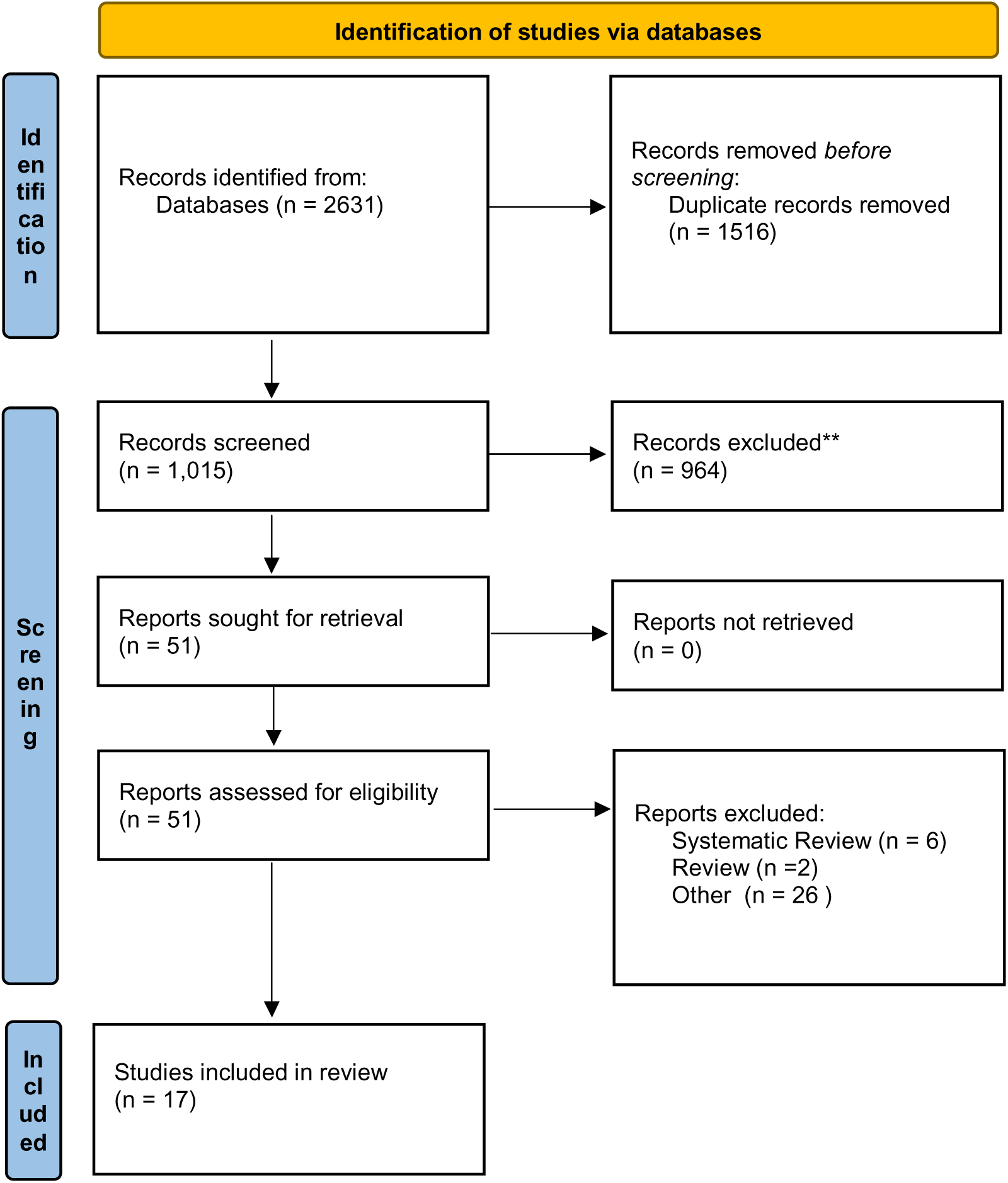

#### Included papers

The first published paper included was by Cantor et al. (1992)[34] who conducted a case-control study of newly diagnosed non-Hodgkin lymphoma (NHL) in 622 white men (cases) compared to 1245 randomly sample, matched controls, who were also white men, in the USA, specifically Iowa and Minnesota. The study measured the risk of NHL associated with farming occupation and specific agricultural exposures to pesticides. Men who ever farmed had a relative increased risk of NHL than non-farmers (OR=1.2, 95% CI: 1.0-1.5) independent of crop or animal types. Men who ever handled glyphosate also showed a slight increased risk of NHL, but the association was not statistically significant (OR=1.1, 95% CI: 0.7-1.9) when adjusted for vital status, age, state, cigarette smoking status, family history of lymphohaemotapoietic cancer, high-risk occupations and high-risk exposures. The corresponding relative risk that I calculated from the data in table 6 was 1.091 (95% CI: 0.69 to 1.72; p=0.7055), and an absolute risk difference (in this case increase) of 0.8% (case risk = 0.098 (9.8%) minus control risk= 0.090 (9%)) and a number needed to harm (NNH) of 122.48. Therefore, with each 123 people exposed to glyphosate, 1 individual would get NHL. This study had several strengths including: that it used a large population-based sample in a farming community, used adjusted analyses, had matched cases and controls, and verified NHL cases. However, the main limitation of this study was that there was low power to assess the risk of NHL with glyphosate with only 26 cases of NHL.

The next published paper is by McDuffie et al. (2001)[8] who conducted a case-control study in six Canadian provinces. The study investigated the associations between exposure to a variety of herbicides and NHL in 517 male cases and 1506 controls. In table 2 they report that the odds of NHL was OR=1.26 (95% CI:0.87-1.81; adjusted for age and province of residence) and an OR=1.20 (95% CI: 0.83-1.74; adjusted for age, province of residence, and various medical variables (see table 2, p1158 [8]) for any exposure to glyphosates (10 hours or more per year). In a separate analysis of exposure days per year to glyphosate for risk of NHL, when compared to the unexposed group, the men with >0 and </= 2 days of exposure had an OR=1.00 (95% CI: 0.63-1.57); and for those with > 2 days of exposure per year had an increased risk of NHL with an OR=2.12 (95% CI: 1.20-3.73) with all analyses being adjusted for age and province of residence. This study had several strengths including: a large multisite sample, confirmation of NHL cases using strong methods, matched controls by age, use of a pilot to refine exposure definitions, verification of pesticide use information in a validation pilot, building on a questionnaire used in prior studies, use of an operational manual and trainings for all sites, adjusted statistical analyses, analyses for any use and a dose response model. Drawbacks include a moderately small number of overall cases.

**Table 2:** Risk of bias assessment of cohort.

| **Risk of Bias Item** | **Study (citation)** |
| --- | --- |
|  | Andreotti 2018<br>[15] |
| Representativeness of exposed cohort | 1 |
| Selection of non-exposed cohort | 1 |
| Ascertainment of exposure | 0 |
| Demonstration that outcome of interest was not present at start of the study | 1 |
| Comparability of the cohorts | 1 |
| Assessment of outcome | 1 |
| Was follow-up long enough for outcomes to occur | 1 |
| Adequacy of follow-up | 0 |
| Total score | 6/8 (75%) |

The next study was by Hardell et al. (2002)[7] who conducted a pooled analysis of two case-control studies in Sweden one of NHL (originally reported in 33) and another on hairy cell leukemia (HCL), (originally reported in 34). The pooled analysis of NHL and HCL was based on 515 cases and 1141 controls, matched for age and country. In the univariate analysis, any exposure to glyphosate increased the risk of NHL and HCL (OR=3.04; 95% CI: 1.08-8.52; 8 exposed cases and 8 controls). After adjusting for study, study area and vital status in the multivariate analysis, the odds of NHL and HCL due to any exposure to glyphosate was OR=1.85 (95% CI: 0.55-6.20). Drawbacks included: small number of NHL cases, lack of data on high and low exposures specifically for glyphosate (in their dose-response analysis they grouped glyphosates with other herbicides and did show increased risk of NHL with increased exposure days, see table 2 [7]). Strengths include the large sample, adjusted analyses and inclusion of deceased individuals so as to not have missing data on exposure (family members were asked about exposures in those that were deceased).

The next study was by, De Roos et al. (2003)[4], which used pooled data from three case-control studies on NHL conducted in the 1980s in Nebraska [37], Kansas [38], and Iowa and Minnesota [34] to examine pesticide exposure in farming as a risk factor for NHL among men.

The three pooled studies all had slightly different methods of study recruitment. The pooled sample population included 870 cases and 2,569 controls – the majority of cases (n=650) and controls (n=1933) were included for the analysis of 47 pesticides controlling for potential confounding by other pesticides, those pesticides for which at least 20 persons were exposed. Logistic regression and hierarchical regression models (adjusting estimates based on prior evidence that the 47 pesticides may cause any type of cancer) were used and all models were adjusted for age, study site, and all other pesticides. In the logistic regression model based on 36 cases the odds ratios for association between any exposure to glyphosate and NHL were 2.1 (95% CI: 1.1-4.0) and in hierarchical regression model the OR=1.6 (95% CI: 0.9-2.8). Of note, the hierarchical regression method (a type of Bayesian analytic technique) has questionable merit here since the adjustments were based on prior evidence of factors that may cause any cancer and not specifically NHL, and the prior evidence used for the adjustments (the priors) was quite dated. This study did have several drawbacks: there was missing data on approximately 25% of the original sample (who answered did not know to exposure on one or more of the 47 pesticides), and who were excluded from the analysis, they did not perform a sensitivity analysis for these participants, but the remaining groups still appeared to be relatively similar. The major strength of this study is the pooled nature of the analysis and the control for other pesticide use in their models.

The next study was by Lee et al. (2004) [39] evaluated whether asthma acts as an effect modifier of the association between pesticide exposure and NHL. The study was conducted using a pooled analysis of population-based case-control studies [37,34] in Iowa, Minnesota and Nebraska, also used in the pooled analysis of De Roos discussed above [4]. The sample included both men and women; 872 cases with NHL from 1980 to 1986 and 2,381 frequency-matched controls. A total of 177 subjects (45 cases, 132 controls) reported having been told by a clinician that they had asthma. The risk of NHL was elevated in both non-asthmatics (OR=1.4, 95% CI: 0.98-2.1) and asthmatics (OR=1.2, 95% CI: 0.4-3.3), though neither OR was statistically significant. This study is redundant with the De Roos study [4] and therefore will not be considered further and is not included in the meta-analyses below.

The next study, by Eriksson et al (2008) [6] reported the results of a case-control study of exposure to pesticides as a risk factor for NHL, with the sample drawn from Sweden. After controlling for age, sex, and year of diagnosis (for cases) or year of enrollment (for controls), which they did in all analyses, the odds of NHL for exposure to glyphosate was 2.02 (95% CI: 1.10-3.71). In their latency analysis, for 1-10 years of exposure the OR for NHL was 1.11 (95% CI: 0.24-5.08) and for >10 years the OR for NHL was 2.26 (95% CI: 1.16-4.40). When considering exposure for more than 10 days per year, the OR was 2.36 (95% CI: 1.04-5.37) and for </= 10 days the OR=1.69 (95% CI: 0.70-4.70). Exposure to glyphosate was associated with increased odds for lymphoma subtypes and elevated odds of B-cell lymphoma (OR=1.87, 95% CI: 0.998-3.51) and the subcategory of small lymphocytic lymphoma/chronic lymphocytic leukemia (OR=3.35, 95% CI: 1.42-7.89). This study has several strengths including: confirmation of cases by independent pathologists, randomly sampled matched controls (for age and sex), blinded interviewers, dose response analyses, latency analyses, and a large sample size. Drawbacks included: 253 cases (over 20% of the original cases) being excluded for various reasons. Overall, this case-control study was well done.

The next study, by Orsi et al. (2009)[9] was a case-control study conducted across six hospitals in France between 2000 and 2004. The study sample included men and women aged 20-75 years and controls that were matched of the same age and sex as the cases were recruited in the same hospital, though only men were included in their analyses. All cases were diagnostically confirmed by a panel of pathologists and hematologists; in-person interviews and expert review of cases were used to evaluate pesticide exposure, with 168 cases being re-interviewed because of insufficient information. They fitted unconditional logistic models, stratifying for age and center, also performed tests for trend of duration and examined the robustness of results using conditional logistic regression restricted to the pair-matched case-control samples. Further, a sensitivity analysis was performed by excluding the subjects in each center and the controls sharing the same broad reason for admission, in turn. The analysis included 491 cases (244 cases of NHL) and 456 controls. For the association between any Glyphosate exposure and NHL, they report an OR=1.0 (95% CI: 0.5-2.2). They also performed an analysis for NHL subtypes, diffuse large cell lymphoma and follicular lymphoma, also finding non-statistically significant ORs (see table 4). The main drawback of this study was the small sample of participants reporting exposure to glyphosate. The main strengths included: Cases recruited within 3 months of diagnoses, matched controls, adjusting for age and center in all analyses, sensitivity analyses, small number of dropouts, blinding of patients and interviewers.

**Table 4:** G.R.A.D.E. summary table.

| Certainty Assessment |  |  |  |  |  | Publication Bias | Overall GRADE | No. of Patients |
| --- | --- | --- | --- | --- | --- | --- | --- | --- |
| *Outcome Measure* | *Study Design* | *Risk of Bias* | *Inconsistency* | *Indirectness* | *Imprecision* |  |  |  |
| **NHL** | All observational | Some but not critical | none | none | Some but not critical | None or unlikely | moderate | 436,545 |

Cocco et al. (2013)[40] reported on a pooled analysis of case-control studies conducted in six countries between 1998-2004 (EPILYMPH, Czech Republic, France, Germany, Ireland, Italy, and Spain) investigating the role of occupational exposure to specific groups of chemicals in the etiology of lymphoma overall, B-cell lymphoma, and several subtypes; they included 2,348 cases and 2,462 controls. The controls for samples from Germany and Italy were randomly selected from the general population and matched on gender, 5-year age-group and residence area; with other countries used matched hospital controls. Participation rate was 88% for the cases, 81% for hospital controls and 52% of population controls participated. In-person interviews, by trained interviewers, were conducted to collect detailed information on occupational history on farm-specific work related to type of crop, farm size, pest being treated, type of schedule of pesticide use. Experts and an agronomist at each study center assessed exposure metrics and a cumulative exposure score was calculated. Unconditional logistic regression models were used (adjusted for age, sex, education, and study center) to estimate the risk of B-cell lymphoma, DLBCL, and CLL associated with ever exposure. The OR for ever-exposure to glyphosate for B-cell lymphoma was 3.1 (95% CI: 0.6-17.1), with 4 exposed cases and 2 exposed controls. Overall, this study had very good methods, as described above, but had a very small number of cases and controls with B-cell lymphoma exposed to glyphosate (2 and 4 respectively). Therefore, the power in this study to detect the association is relatively low, as reflected in the very wide confidence intervals.

The next published study was by Andreotti (2018) [17], an update of the prospective cohort study reported by De Roos (2005)[5], from the study called the Agricultural Health Study (AHS) which includes licensed pesticide applicators from North Carolina and Iowa. Between 1993 and 1997 57,310 individuals were enrolled, with 63% completing follow-up phone interviews 5 years after enrollment (1999-2005). Cancer diagnoses were done via registry linkage, classified according to the ICD, 3^rd^ revision, and vital status was determined through state death registries and the National Death Index. A questionnaire was used to determine glyphosate and other pesticide exposure, both at enrollment and at each follow-up (e.g., number of days each pesticide was used) and several lifetime exposure metrics were created including ever/never use, lifetime days of use, and intensity-weighted lifetime days (intensity days x intensity score. The intensity score was determined from literature-based measurements and information provided by the applicator. Multiple imputation was used to impute missing data. After exclusions, 54,251 applicators were enrolled but only 63% completed follow-up phone interviews 5 years later. Poisson regression was used to estimate incidence rate ratios and 95% confidence intervals; cumulative lifetime days and intensity-weighted lifetime days were separated into quartiles, tertiles, or medians; risk estimates were adjusted for age, cigarette smoking, alcoholic drinks, family history of cancer, state, and the 5 pesticides most highly correlated with glyphosate. They also evaluated potential confounding factors (e.g. BMI and pack-years of cigarette smoking) and adjusted for solvents linked to cancer risk. Finally, they conducted sensitivity analyses using additional exposure information. The authors report, in their primary analysis, that for the unlagged intensity-weighted lifetime days of glyphosate exposure, there were no statistically significant associations between glyphosate and NHL or any NHL subtypes. They report for NHL and the top exposure quartile a RR =0.87 (95%CI: 0.64-1.20, p=0.95), which did not change when MM was excluded. The findings were unchanged in the sensitivity analyses including further adjustment for confounders. In the supplementary data which report the cancer incidence for lifetime days of glyphosate exposure they also showed no significant relative risk trends across quartiles of exposure for NHL (p=0.44) though they did not do any analyses on the any lifetime exposure. Furthermore, the lagged exposure data in table 2 of the supplementary material show no associations for glyphosate exposure and NHL at 10 or 15 years, for trends across quartiles, but again no any lifetime use analyses were shown. This study has several strengths including: its prospective design, large sample size and event rate, controlling for confounds and sensitivity analyses. But this study had several major drawbacks including: a 37% loss to follow-up, questionable method of imputation [41] and the lack of any lifetime use analyses.

The next study was my Leon et al (2019)[20] which was a pooled analysis of 3 large agricultural worker cohorts, the Agricultural and Cancer (AGRICAN) cohort from France [42], the Cancer in the Norwegian Agricultural population (CNAP) cohort [43], and the AHS cohort [44] from the USA, the latter as reported in Andreotti above. This study was done within the AGRICOH consortium [45] and looked at the risk of NHL and major subtypes in 316,270 farmers and farm workers. The cohorts were included that met certain conditions including periodic data linkage to cancer incidence registries, availability of data on pesticides and or crop cultivation to estimate exposure and sample sizes that were large enough to study NMH subtypes. The harmonizing of pesticide exposure data across the cohorts was published elsewhere [46]. They looked at fourteen chemical groups. They developed crop-exposure matrices (CEMs) for each cohort, described assignment of potential exposure, and the main endpoint was first incident NHL during follow-up, with follow-up time being calculated as number of days between start of follow-up and first date of incident cancer, loss to follow-up or migration out of the area, death or end of follow-up. Statistical analyses included imputation for missing data, cox proportional hazards models, with all covariates being time dependent, and all models used age at date of censoring as the time scale and were first adjusted for sex and animal production, and other covariates included. Fully adjusted models were built for each cohort separately. Models were run for each cohort individually and combined using a random effects meta-analysis (though they do not state what exact model) and I^2^ was calculated to express the magnitude of statistical heterogeneity across cohorts. They report a HR of having NHL in those ever exposed to glyphosate as (95% CI: 0.77-1.18) with an I^2^=57% and p=0.10, for CLL/small lymphocytic lymphoma a HR of 0.92 (95% CI: 0.69-1.24), I^2^=0%, p=0.38, for DLBCL a HR=1.36 (95% CI: 1.00-1.85), I^2^=0%, p=0.48, for follicular lymphoma HR=0.79 (95%CI: 0.52-1.21), I2=0% p=0.56, and for MM a HR=0.86 (95% CI: 0.66, 1.15), I^2^=0%, p=0.95. This pooled analysis, an individual patient data meta-analysis, has several strengths including: Its large size and the use of data from prospective cohort studies. The drawbacks in this study include: the lack of details of the meta-analytic procedures, the lack of assessment of publication bias, a questionable imputation method for AGRICAN and AHS data [54], and potential for exposure misclassification.

The next study is by Pahwa (2019)[23] was a pooled analysis of case-control studies in the US and Canada [8,34,37,38], with Cantor [34] and McDuffie [8] being described above. This pooled analysis used the original histology codes to classify NHL cases using the ICD-0-1 scheme, they estimated several exposure metrics, used simple (and questionable) imputation method for missing data, used unconditional logistic regression for NHL overall and several NHL subtypes controlling for age, state/province, sex, lymphatic or hematopoietic cancer in a first relative, response by a proxy and use of personal protective equipment (PPE) as well as farming and medical factors, the latter two of which was not retained in the final model due to lack of impact on the OR. They also looked at the possibility of confounding by pesticide use.

They investigated trends for the duration, frequency and lifetime days of glyphosate use and NHL and sensitivity analyses were done excluding proxy respondents from the main analysis. The study included 1690 NHL cases and 5131 controls, with all being included in the ever/never analysis, and smaller sample sizes being included in the duration of use and frequency and lifetime days analyses. The authors report a significant association between ever use of glyphosate and NHL overall in the first adjusted model (adjusted for age, state/province, sex, lymphatic or hematopoietic cancer in a first relative, response by a proxy and use of personal protective equipment (PPE)) without controlling for other pesticide use with an OR = 1.43 (95% CI: 1.11-1.83), but the relationship decreased after additionally adjusting for 2 pesticides, resulting in an OR of 1.13 (95%CI: 0.84-1.51). Also, the OR for the DLBCL was initially statistically significant in the first adjusted model but not in the other pesticide adjusted model, the same results being found for other NHL subtypes. In the analysis for number of days per year exposed to glyphosate, individuals who handled glyphosate for >2 days per year had an OR of 2.42 (95% CI 1.48-3.96) for NHL overall in the first adjusted model (double the OR compared to no exposure or >0 to </= 2 days per year) and this statistically significant relationship persisted after controlling for the use of 2,4-D, dicamba and malathion (or=1.73, 95% CI: 1.02-2.94), with similar results for DLBCL but not for other subtypes (see table 4 in Pahwa). They also report a statistically significant trend in overall NHL risk for lifetime days of use of glyphosate (p=0.05) with ORs going from 1.0 to 1.20 to 1.55 in the first adjusted model but not in the other pesticide additionally adjusted model. The sensitivity analyses had no impact on the overall findings and there did not appear to be any between study heterogeneity. Overall, this study was very well done with strengths including: pooled analyses, sensitivity assessments, dose and exposure responses assessments, a large sample size, adjustment for other pesticide use.

The next included study, published in April of 2021 by Meloni et al [47] was a multi-centered case-control study conducted in Italy, between 2011 and 2017, across six centers including 867 lymphoma cases and 774 controls, the latter being hospital-based cancer free from surgery wards, eye care departments, hematology outpatients or patients suffering from traumatic injuries, gastrointestinal disorders, cardiovascular disorders. At recruitment 7.4% of cases and 38.4% of controls refused to participate. The investigators used a detailed questionnaire given by trained interviewers, and occupational physicians and industrial hygienists, with expertise in the retrospective assessment of agricultural exposures, and assessed exposure to glyphosate classified by duration, confidence, frequency, and intensity of exposure to glyphosate for each study subject. They classified lymphoma according to the 2008 update of the WHO classification of lymphoma, which relies on morphology, immunophenotype, molecular biology, genetics, and clinical presentation and course of the disease, and it includes all the lymphoma subtypes originating from B-lymphocytes, including CLL and MM, among the group of B-cell lymphoma (BCL). T-cell lymphomas, and Hodgkin lymphoma (HL) were separately classified. They then used unconditional regression analysis, and modelled risk of major lymphoma subtypes associated with exposure to glyphosate and adjusted for age, gender, education, and study center. They report an OR of 1.4 (95% CI: 0.62-2.94) for NHL and ever glyphosate exposure and an OR of 1.2 (95%CI: 0.35-4.21) for those ever exposed with medium-high confidence. They also report for B-cell lymphoma (B-CL) and OR of 1.6 (95% CI: 0.73-3.66), for DLBCL an OR=0.80 (95%CI: 0.10-6.62), for Follicular lymphoma (FL) an OR=3.7 (95%CI: 1.06-12.8) and for Chronic Lymphocytic Leukemia (CLL) an OR=0.6 (95% CI: 0.11-3.48) in those ever exposed.

For those ever exposed with medium-high confidence the former 2 ORs for B-CL and DLBCL remained similar but for FL the OR went up to 7.1 (95% CI=1.57-31.9). In their analysis in relation to exposure metrics, they reported several statistically significant associations for duration, intensity, frequency and cumulative exposure (see table 4 in Meloni et al [45]). This study had several strengths including: a decently large sample, multicentered design, careful exposure determination, adjusted analyses. But this study also had several drawbacks; small number of cases (lack of power to detect small associations), no sensitivity analyses, lack of control for other pesticides, lack of known reasons for refusal for involvement at recruitment, and lack of details of Lyphoma diagnostic methods or confirmation.

The next study was by Poh et al [57] which was a population-based study conducted using the California cancer registry. They identified patients with a first primary diagnosis of NHL from 2010 to 2016 and linked these patients with CalEnviroScreen 3.0 to obtain production agriculture pesticide exposure to 70 chemicals from the state-mandated Pesticide Use Reporting (PUR) by census tract from 2012 to 2014. In addition, they took data from PUR and integrated it into a geographic information system that employs land-use data to estimate cumulative exposure to specific pesticides previously associated with NHL (glyphosate, organophosphorus, carbamate, phenoxyherbicide, and 2,4-dimethylamine salt) between 10 years prior up to 1 year after NHL diagnosis. They used Multivariable Cox proportional hazards regression models to evaluate the association between total pesticide exposure from CalEnviroScreen 3.0 and individual pesticide exposure from geographic land use data and lymphoma-specific and overall survival. Models included variables such as: gender, race/ethnicity, age and stage at diagnosis, modality of initial therapy, health insurance status, neighborhood SES and rural or urban MSSA. They included 35,808 NHL patients with 44.2% being exposed to a pesticide in their census tract of residence. Glyphosate exposure was observed in 34.1%, of NHL patients. They report a HR of

1.01 (95% CI 0.96-1.06) for lymphoma-specific or overall survival in NHL patients exposed to Glyphosate. They also separated total exposure into low, mid and high exposures reporting HR of 1.02 (95% CI: 0.96-1.09), 1.04 (95% CI: 0.98-1.11) and 1.02 (95% CI: 0.95-1.08). Furthermore they report HRs for NHL histologic subtypes in their table 3.

**Table 3:**
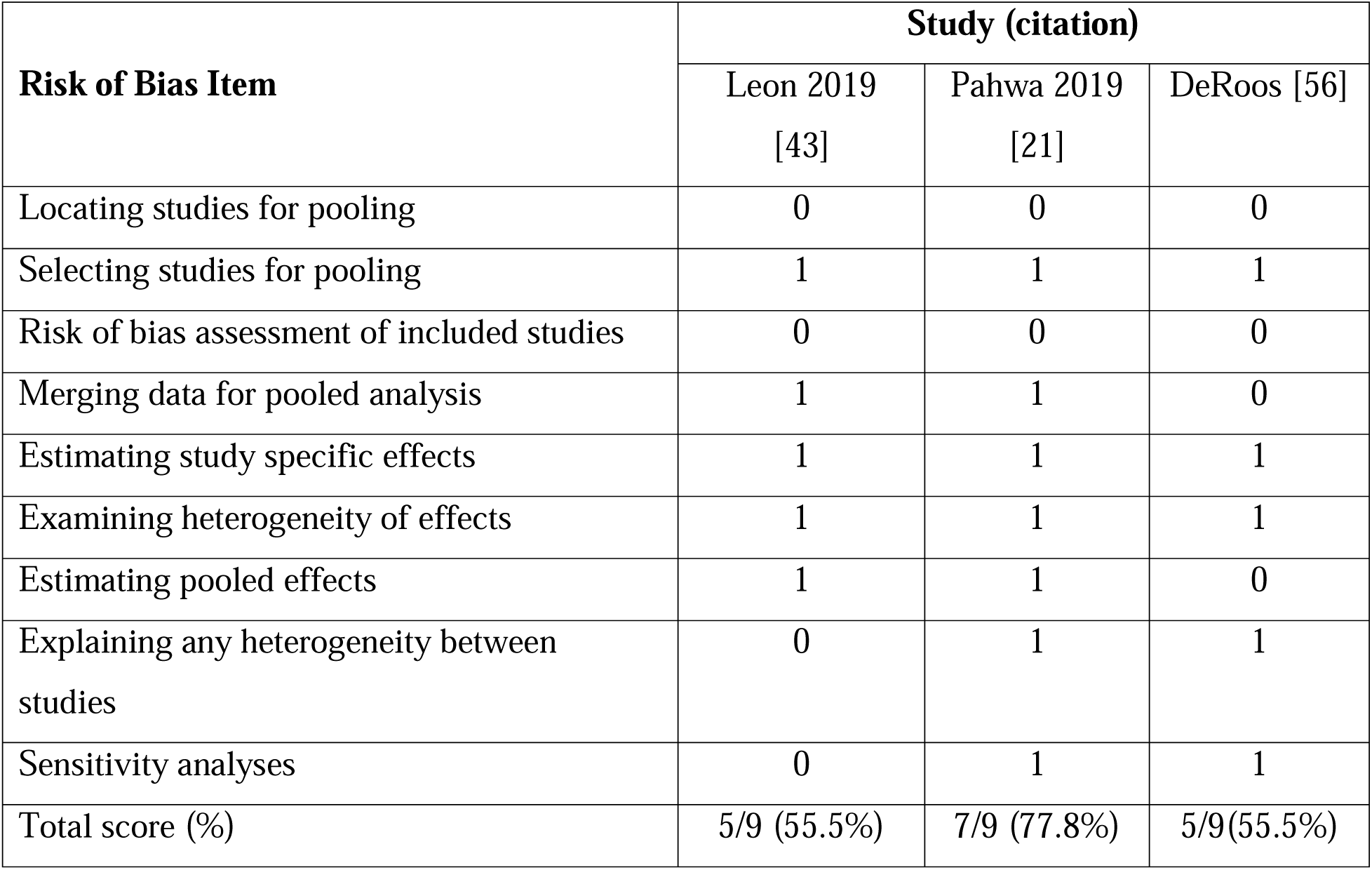
Risk of bias assessment of pooled analyses.

The next and most recent study by De Roos et al (2022) [58] was a pooled analysis of 10 separate data sets, two of which were Cocco 2013 and Orsi 2009, described above. This study pooled data from 10 case-control studies participating in the International Lymphoma Epidemiology Consortium, including 9229 cases and 9626 controls from North America, the European Union and Australia. Herbicide use was coded from self-report or by expert assessment in the individual studies, for herbicide groups (eg, phenoxy herbicides) and active ingredients (eg, 2,4-dichlorophenoxyacetic acid (2,4-D), glyphosate). The authors assessed the association between each herbicide and NHL risk was estimated using logistic regression with adjustment for sociodemographic factors, farming and other pesticides. A total of 8 of these studies assessed the glyphosate exposure and NHL relationship. The authors report an association between glyphosate and follicular lymphoma (lagged 10 years: OR=1.48, 95% CI: 0.98 to 2.25). The authors include multiple myeloma (MM) in their definition of NHL, which is not consistent with current classification systems. When MM is taking out of the OR calculation from table 2, this results in an OR=1.13 (95% CI: 0.97-1.33, p=0.108) for risk of any type of NHL in those ever exposed to glyphosate. This study is difficult to properly assess methodologically because the details of the methods of all data sets from the 8 included studies are not reported and not available elsewhere.

#### Risk of Bias Assessment

The risk of bias assessment for each study is listed in table 1,2 and 3 below. Table 1 includes risk of bias assessments for case-control studies, table 2 for cohort studies and table 3 for pooled analyses. Overall, there were 15 disagreements between assessors, out of a total of 71 total assessments, all of which were resolved easily by discussion and review of the articles.

While we included 17 separate studies from our review of the peer-reviewed literature, several of these overlapped either partially or fully with other studies that pooled multiple individual studies [20,23,58], the latter of which are pooled analyses to be assessed for risk of bias separately. Therefore, we assess 10 studies here for risk of bias. Five studies were considered at low risk of bias (>70% of items fulfilled), four studies of moderate risk of bias (between 40 and 69% of items fulfilled) and one study at a high risk of bias (under 40% of items fulfilled).

**Table 1:** Risk of Bias Assessment of Case-Control Studies.

| **Risk of Bias Item** | **Study [citation]** |  |  |  |  |  |
| --- | --- | --- | --- | --- | --- | --- |
|  | Hardell<br>2002 [5] | Eriksson<br>2008 [4] | Orsi<br>2009 [7] | Cocco<br>2013 [8] | Meloni<br>2021<br>[45] | Poh [55]^ |
| Adequate Case Definition | 1 | 1 | 1 | 1 | 0 | 1 |
| Representativeness of Cases | 1 | 1 | 1 | 1 | 0 | 1 |
| Control Selection | 1 | 1 | 0 | 0 | 0 | 0 |
| Definition of Controls | 0 | 0 | 1 | 1 | 0 | 0 |
| Controls for Other Pesticides | 1 | 1 | 0 | 0 | 0 | 0 |
| Controls for Age | 1 | 1 | 1 | 1 | 1 | 1 |
| Exposure Assessment | 1 | 1 | 1 | 0 | 0 | 1 |
| Method Consistency | 1 | 1 | 1 | 1 | 1 | 1 |
| Non-Response Rate | 1 | 1 | 1 | 0 | 0 | 1 |
| Total score | 8/9<br>(88.9%) | 8/9<br>(88.9%) | 7/9<br>(77.8%) | 5/9<br>(55.9%) | 2/9<br>(22.2%) | 6/9(66.6%) |

Poh [55] was a population based study.

#### Meta-analytic Findings

##### Effect estimate choice from each included study

The studies included in the meta-analyses below frequently reported varying effect estimates for example for varying statistical adjustments or not (e.g.,6,23). For each analysis below, we note which effect estimates were used from individual studies. In all cases, where available, we used the reported adjusted analyses from each study, unless those adjusted analyses were under powered, at a high risk of type 1 errors. Regression analyses that include a large number of variables in their adjustments relative to the number of events (e.g., NHL) in the groups being compared (for example exposed vs non-exposed groups) run the risk of being subject to spurious results, they are overfitted. For example, in logistic regression empirical evidence shows that having less than 10 events (e.g. NHL events) per variable in the regression model increases the type 1 error rate [50–52] and more recent evidence shows that even at 20 events per variable, power is very low [53]. Therefore, when the adjusted model was underpowered, for example in Eriksson [6], we used the unadjusted value. Another example of an underpowered adjusted value was from Pahwa [23], in table 2 they report two adjusted analyses for NHL overall for ever glyphosate use. They have an ever-exposed sample of 113 for NHL overall, which is the number of events. In model A they include 6 variables and in model B they include 8 variables. Model A, even at 20 events per variable is underpowered and model B is underpowered to a greater extent, therefore we used the estimate from model A to lessen the risk of spurious findings, though not eliminate them.

##### Ever Exposure Main analyses

The first meta-analysis we ran was a fixed effects inverse variance model using the natural log of the OR and CIs for all included papers, except for Cantor 1992 [34], McDuffie 2001, DeRoos 2003, Lee 2004, since they were all included in the recent follow-up pooled analyses by Pahwa 2019. We also excluded Cocco 2013 and Orsi 2009 since there were included in the recent pooled analyses by DeRoos 2022 [38]. Furthermore, we excluded the study by Hardell (2023) as it included Hardell 1999, Eriksson 2008 which we had already included, and they also included Nordstrom 1998, which focussed on hairy cell leukemia, the latter of which is not typically considered a type of NHL. Therefore, we included 9 published papers that represented 20 unique studies or data sets.

In the fixed effects model the overall meta-analytic OR was 1.033 (CI: 0.988-1.080) with a p= 0.152 but also with a significant test for statistical heterogeneity p=0.028, and an I^2^ value of 53.4%, which represents the existence of heterogeneity. Given the significant test for statistical heterogeneity, and that fact that most studies adjusted their estimates for multiple variables, and that we could not determine variables to explain this heterogeneity, we also ran an inverse variance random effects model which incorporates the heterogeneity into the model which resulted in vary similar findings (see figure 1). In the random effects model the OR=1.114 (0.981-1.273; p=0.141; Figures 1 and 2).

**Figure 1:**
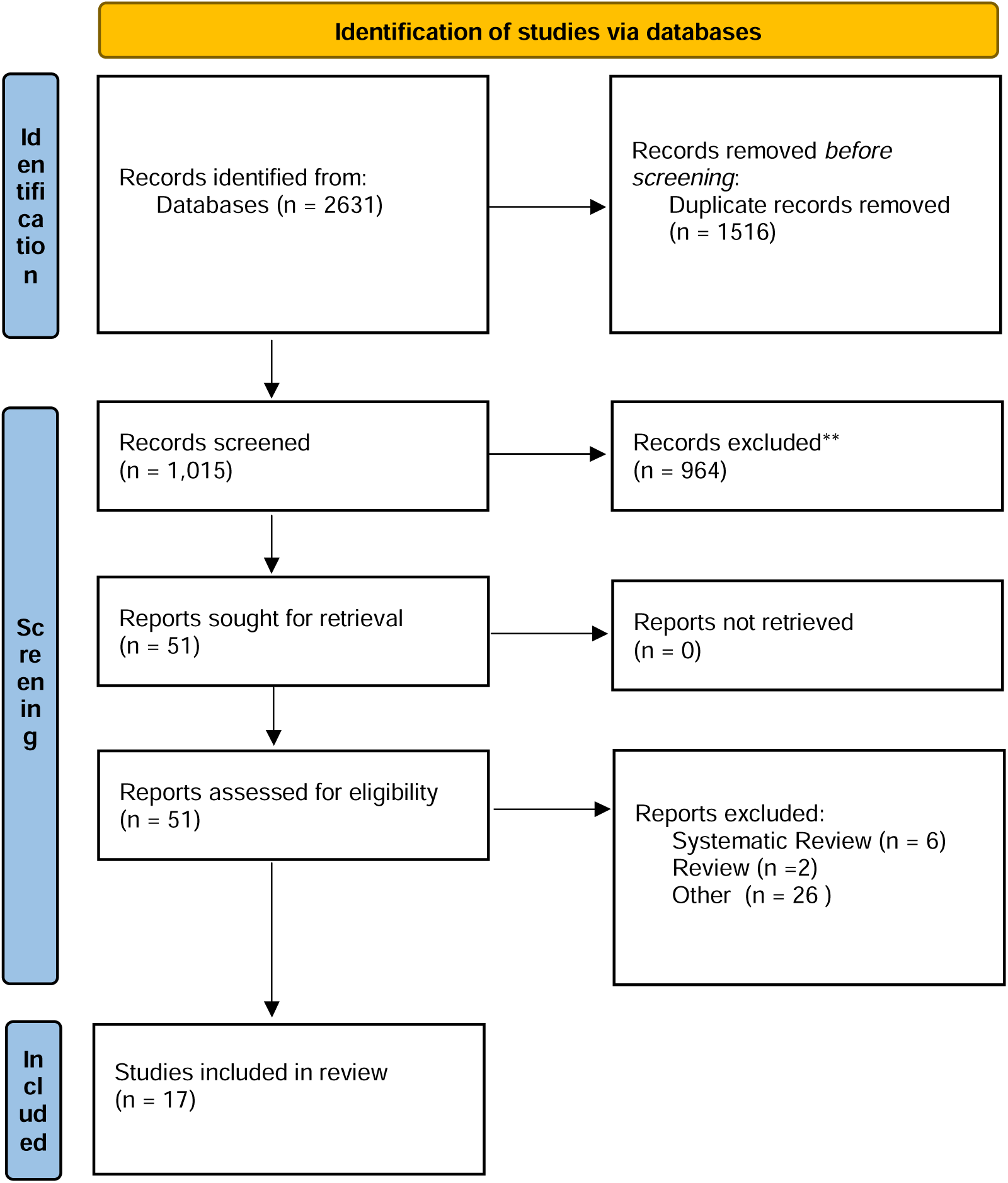
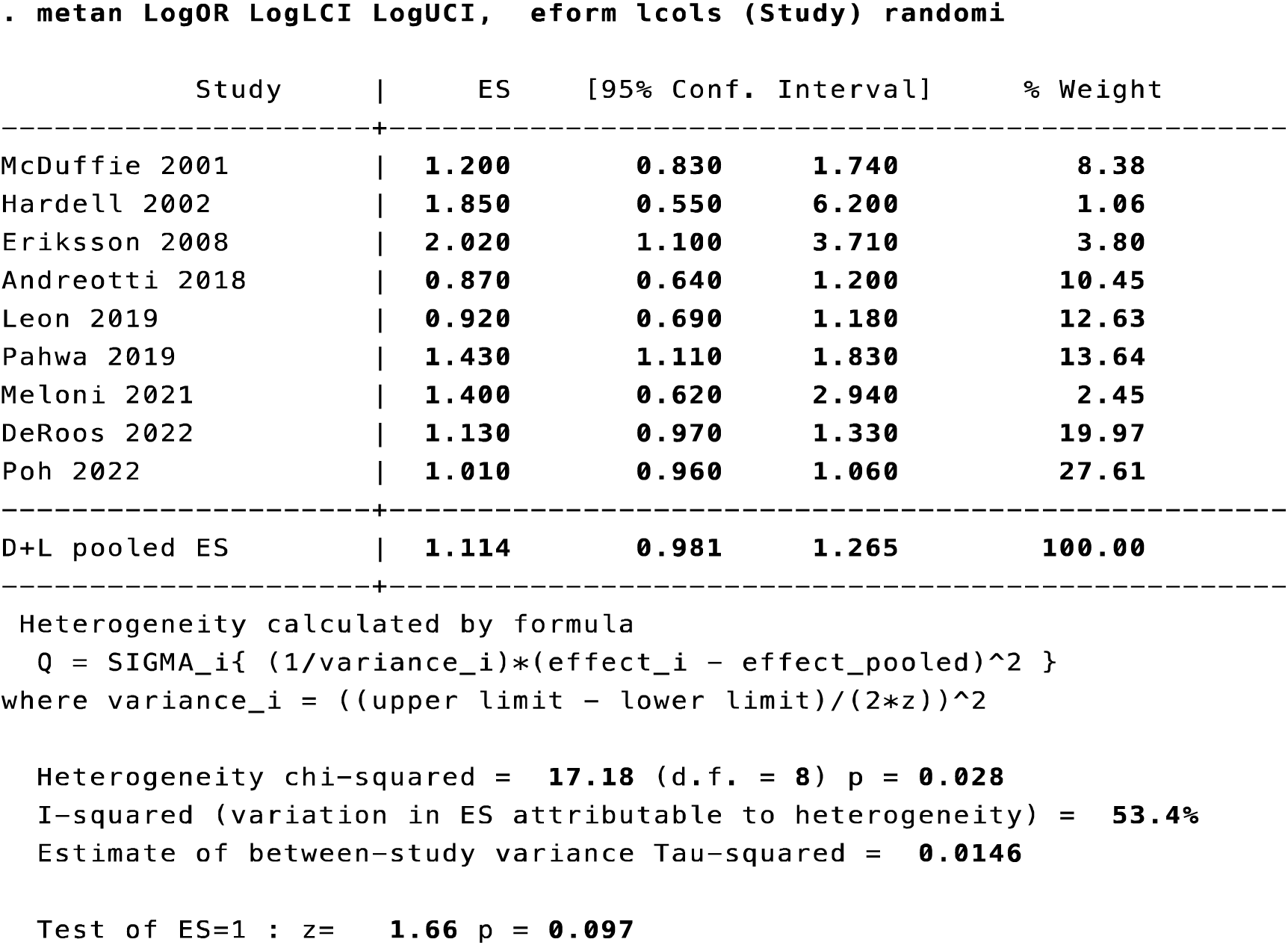
Random Effects Meta-Analysis Results Table for ever exposure

**Figure 2:**
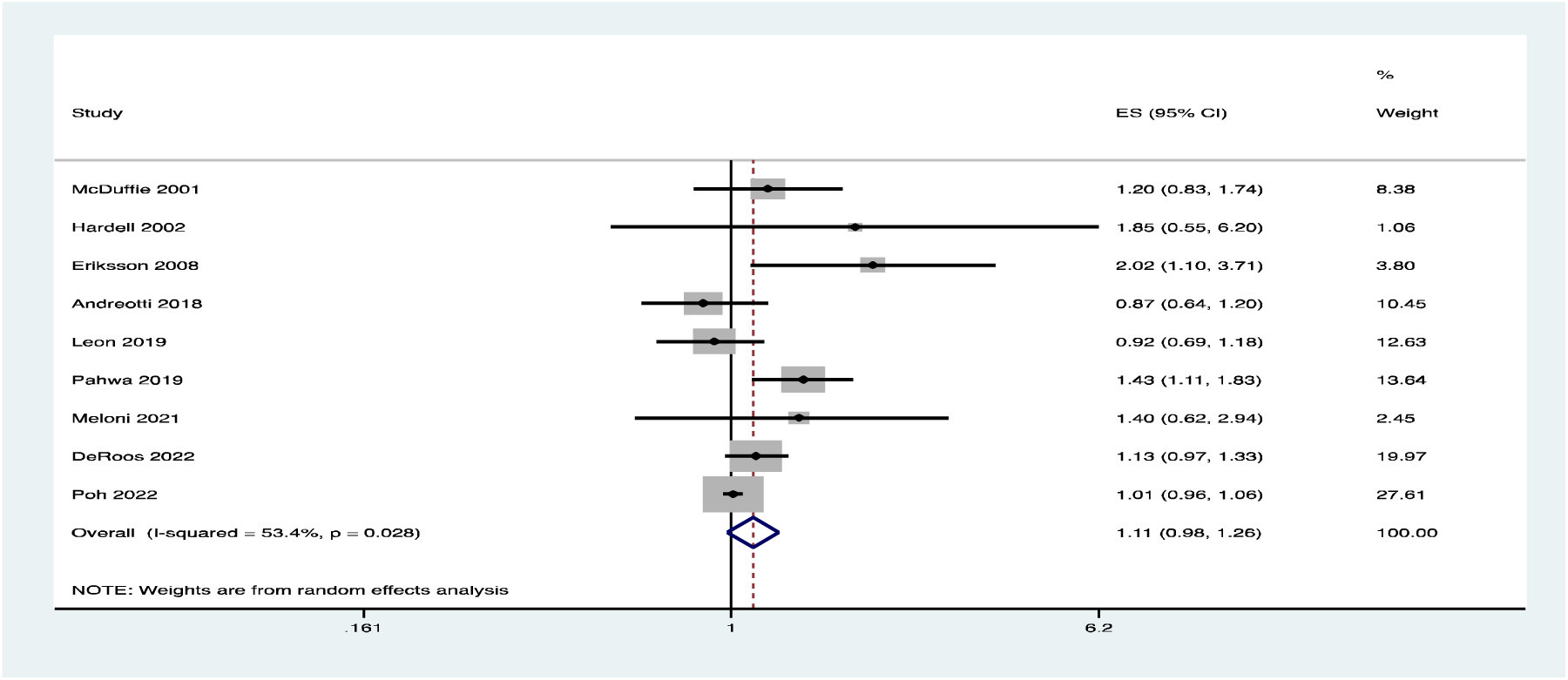
Forest plot for ever exposure across all included studies using random effects model.

We ran another analysis removing Leon and Poh, using a fixed effects model resulting a statistically significant OR=1.185 (95%CI: 1.053-1.334; p=0.001), with an acceptable level of statistically heterogeneity (49.4%; p=0.079). While the amount of heterogeneity was low the p value for the cutoff of statistically significant heterogeneity exceeded the standard accepted p=0.10 for Cochran’s Q test. Therefore, we ran a random effects model that resulted in very similar findings, OR=1.234 (95%CI: 0.996-1.528; p=0.054), with one of the wider confidence intervals. These latter two models are the most reliable models, including the largest number of data sets (n=18) excluding HR estimates (Poh and Leon) and including the most recent AHS data Andreotti (see figures 3 and 4). These analyses also resulted in little heterogeneity using the fixed effects model, though not negligible heterogeneity, with a random effects model yielding very similar results. The results from these analyses suggest an increased odds of risk of NHL (18.5%-24%) in those exposed to Glyphosate at any point in their life compared to those who were not.

**Figure 3:**
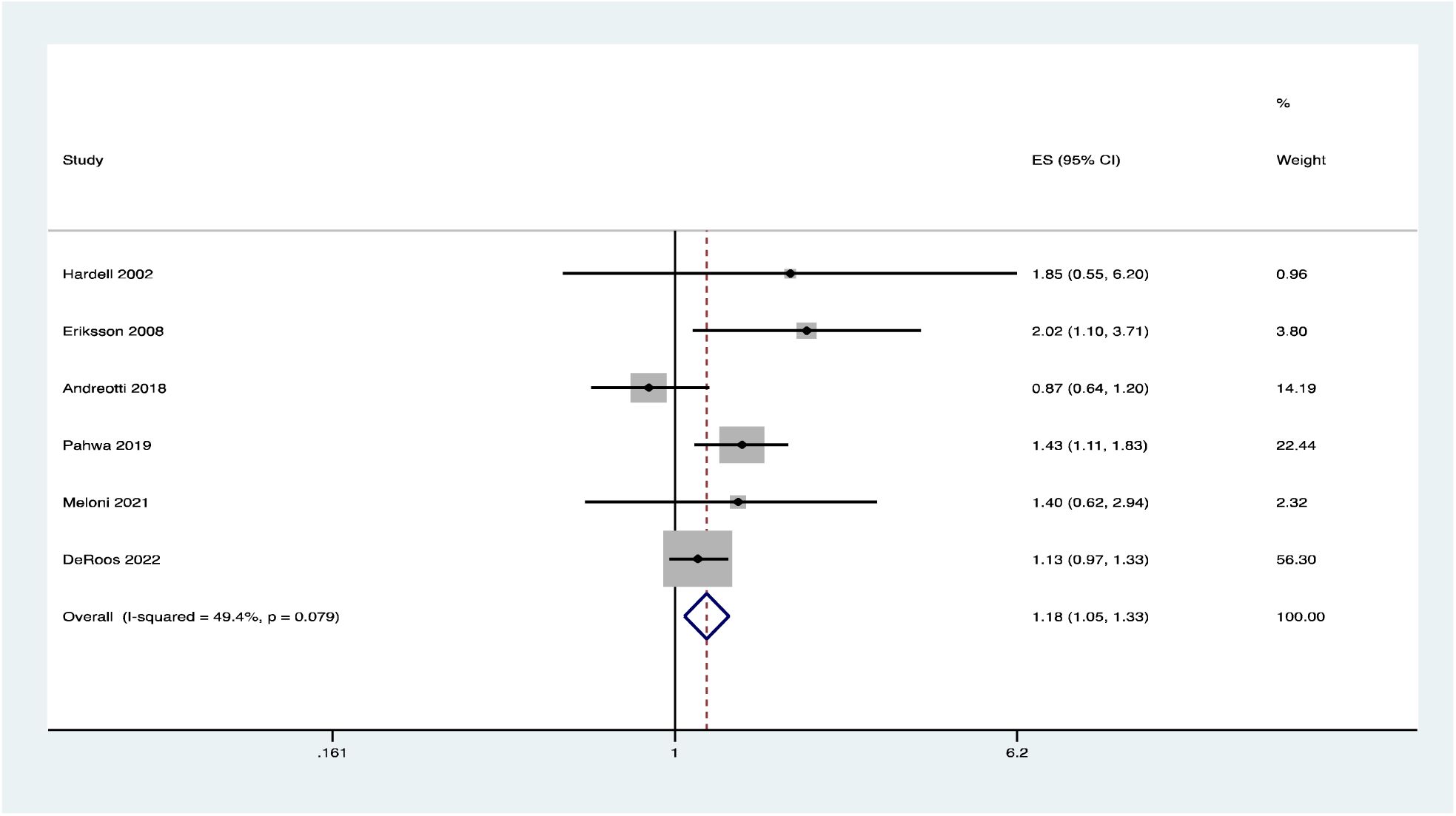
Ever exposure data removing Leon and Poh: Fixed Effects Forest Plot

##### Highest Reported Level of Exposure Meta-Analyses

First, we performed an inverse variance fixed effects model across all studies and found a meta-analytic estimate OR=1.030 (CI: 0.96-1.105), p=0.405 and a statistically significant test for heterogeneity p=0.013 and a moderately high magnitude of statistical heterogeneity I^2^=60.7%.

Given the significant test for statistical heterogeneity, that most studies presented adjusted ORs, and that fact that we could not determine variables to explain this heterogeneity, we also ran an inverse variance random effects model which incorporates the heterogeneity into the model which resulted in a meta-analytic OR=1.183 (95% CI=0.971-1.441), p=0.095 (see figures 5 and 6).

**Figure 4:**
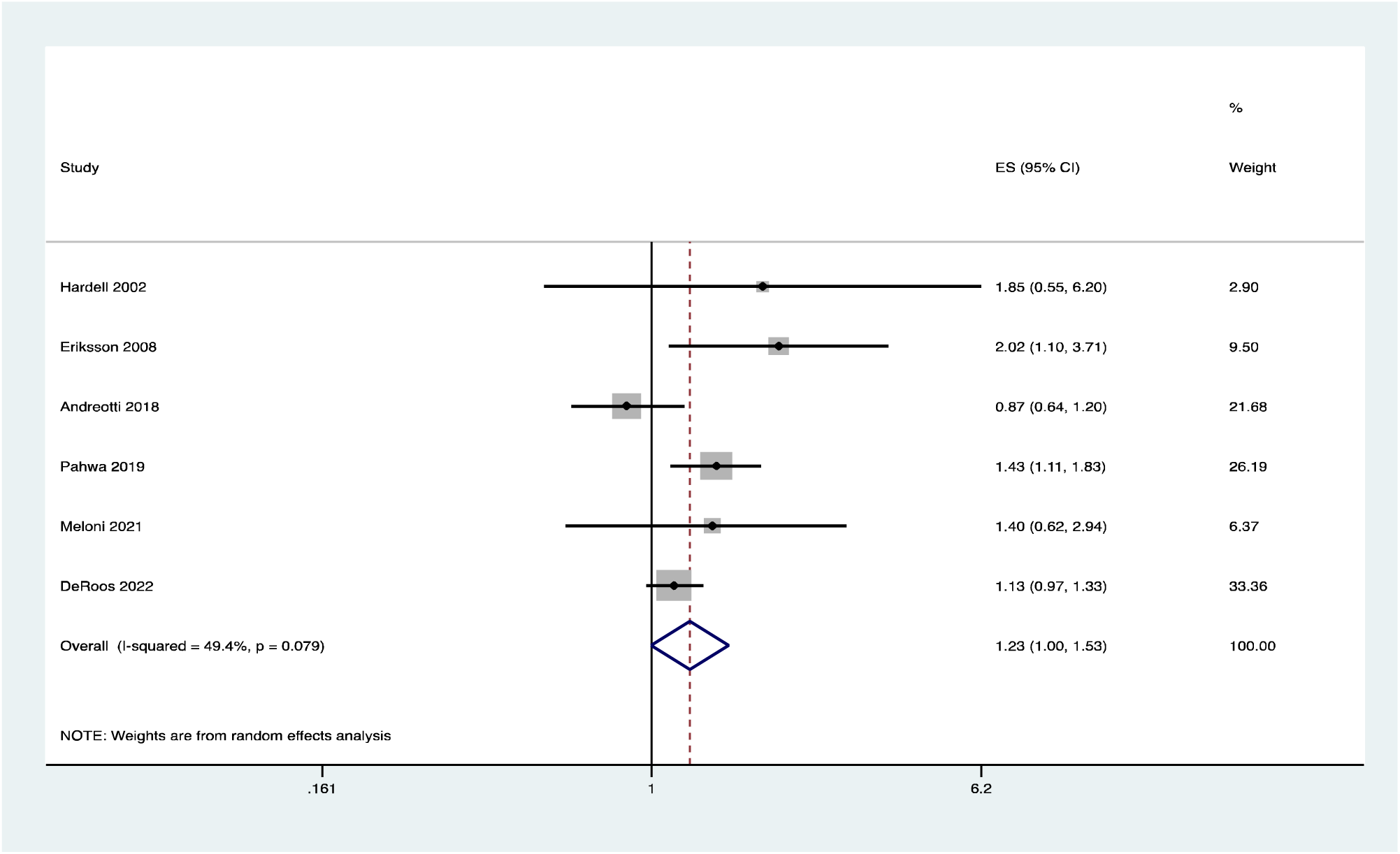
Ever exposure data removing Leon and Poh: Random Effects Forest Plot

**Figure 5:**
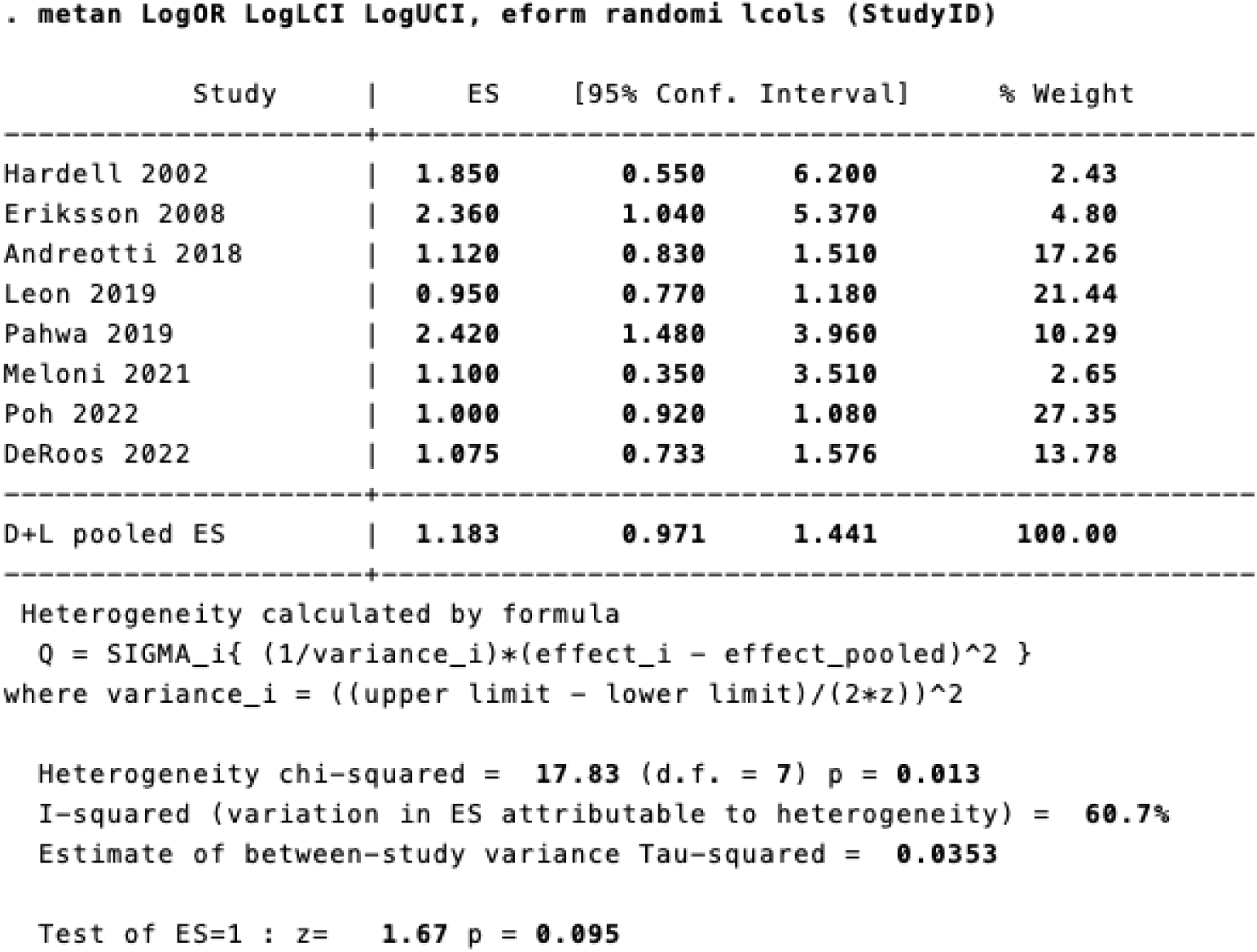
Random Effects Meta-Analysis Results Table for Highest Exposure Category

**Figure 6:**
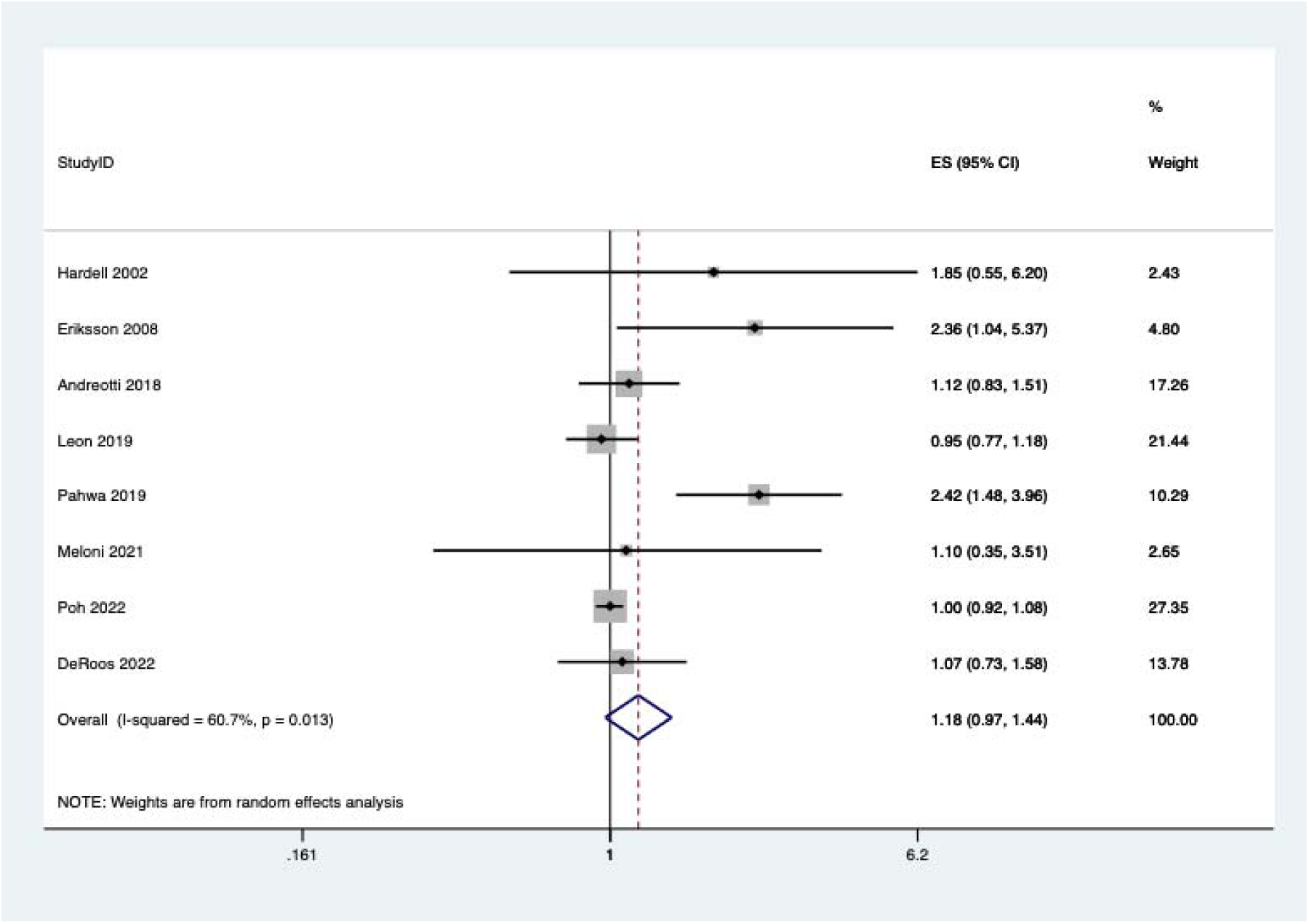
Forest Plot Random Effects Meta-Analysis for highest exposure category

We then did another sensitivity analysis removing Leon and Poh, given that they included HRs only. This resulted in a pooled effect estimate of OR=1.332 (95%CI: 1.090-1.626) p=0.005 with significant heterogeneity at I^2^=52%, p=0.064. Therefore, we ran a random effects model to incorporating the statistical heterogeneity which resulted in meta-analytic OR=1.465 (95%CI=1.043-2.057), p=0.027 (see figures 7 and 8).

**Figure 7:**
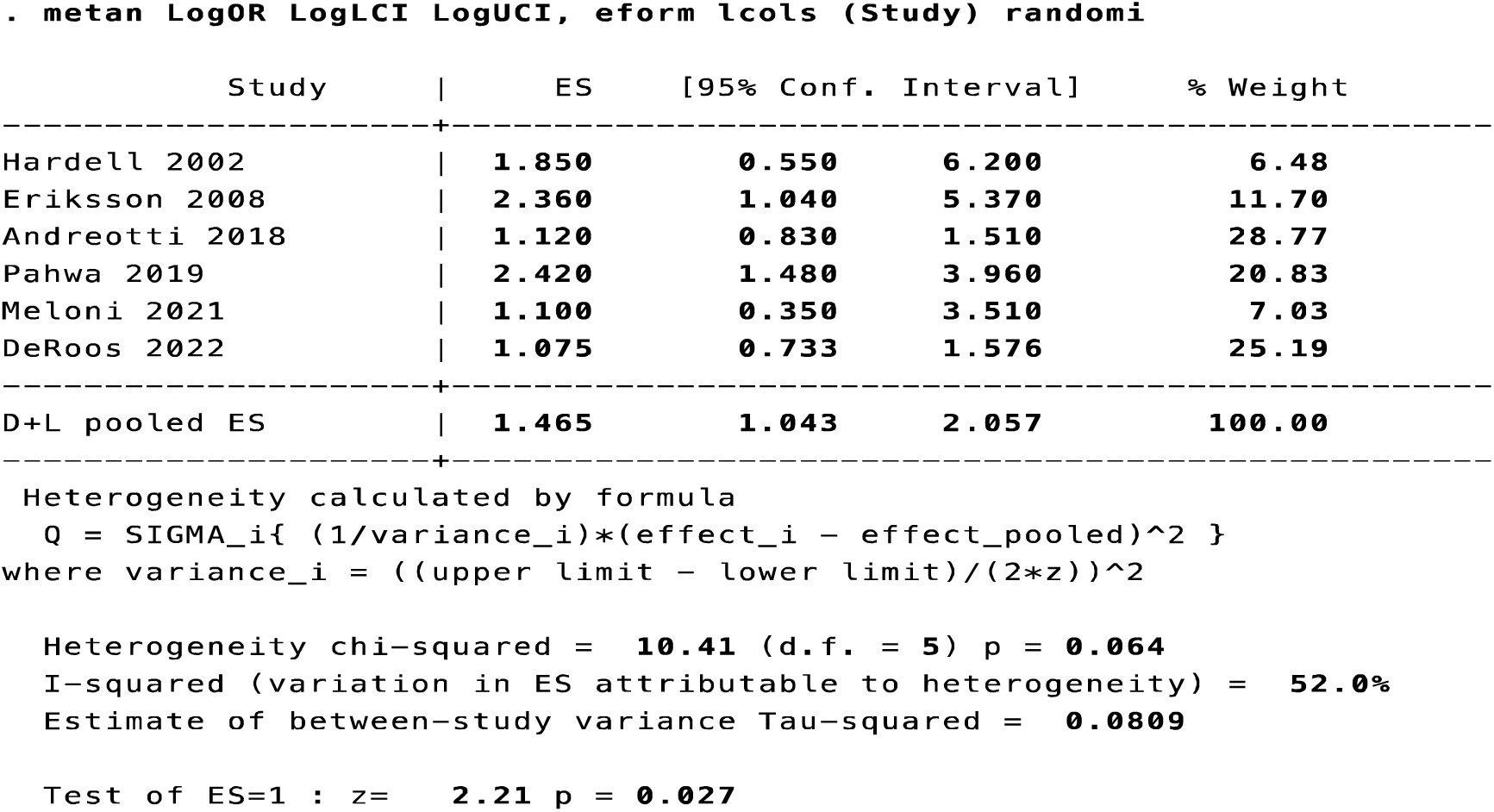
Random Effects Meta-Analysis Results Table for Highest Exposure Category removing Leon and Poh

**Figure 8:**
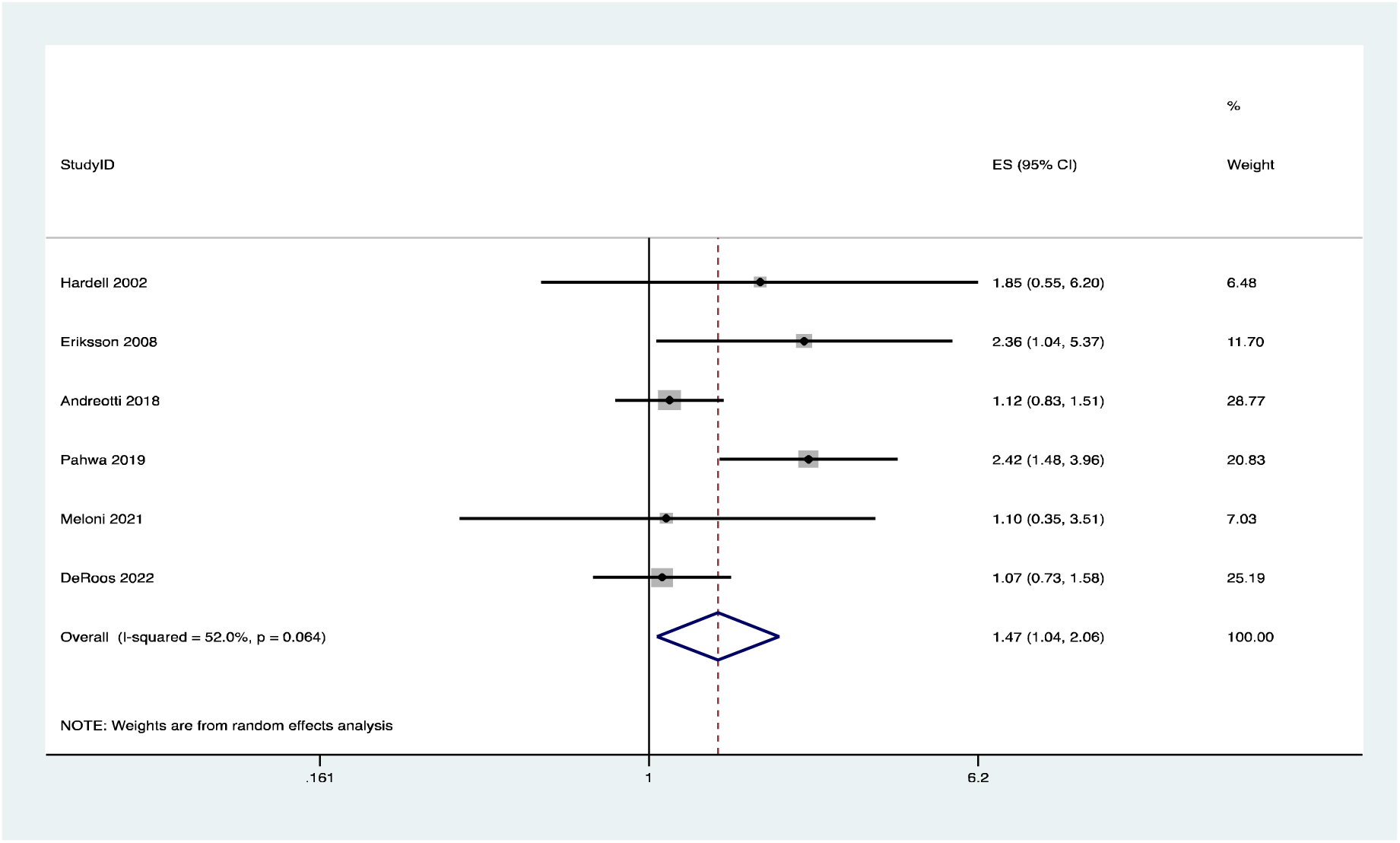
Forest Plot Random Effects Meta-Analysis for highest exposure category removing Leon and Poh

We then ran a sensitivity analysis for my choice of effect estimate from DeRoos 2022. In the highest exposure analysis above (Figure 7 & 8) we used the highest category of exposure from Table 3 in DeRoos (>15.5 years), removing those with multiple myeloma, which is not considered a type of NHL, which resulted in the OR we calculated of 1.075 (CI: 0.733-1.576). We then did a sensitivity analysis and combined the latter two categories in that table, to get an estimate of all those with NHL with >8 years exposure, in which case the OR went up, to 1.117 (CI: 0.866-1.441). We then performed a fixed effects meta-analysis resulting in meta-analytic estimate of 1.28 (CI: 1.08-1.53), p=0.005, with moderately high heterogeneity (p=0.054) thus we ran a random effects meta-analysis resulting in a OR=1.45 (CI: 1.056-1.98), p=0.021. Next, we used the cumulative exposure data from Meloni [45] instead of the frequency of exposure data and the cumulative data from Pahwa, excluding Leon and Poh. This resulted in meta-analytic estimate of OR=1.327 (CI: 1.037-1.697), p=0.024, with a statistically non-significant test for heterogeneity p=0.498 and an I^2^ of 0% (figures 9 and 10). Therefore, across all sensitivity analyses, the results remained relatively consistent and statistically significant.

**Figure 9:**
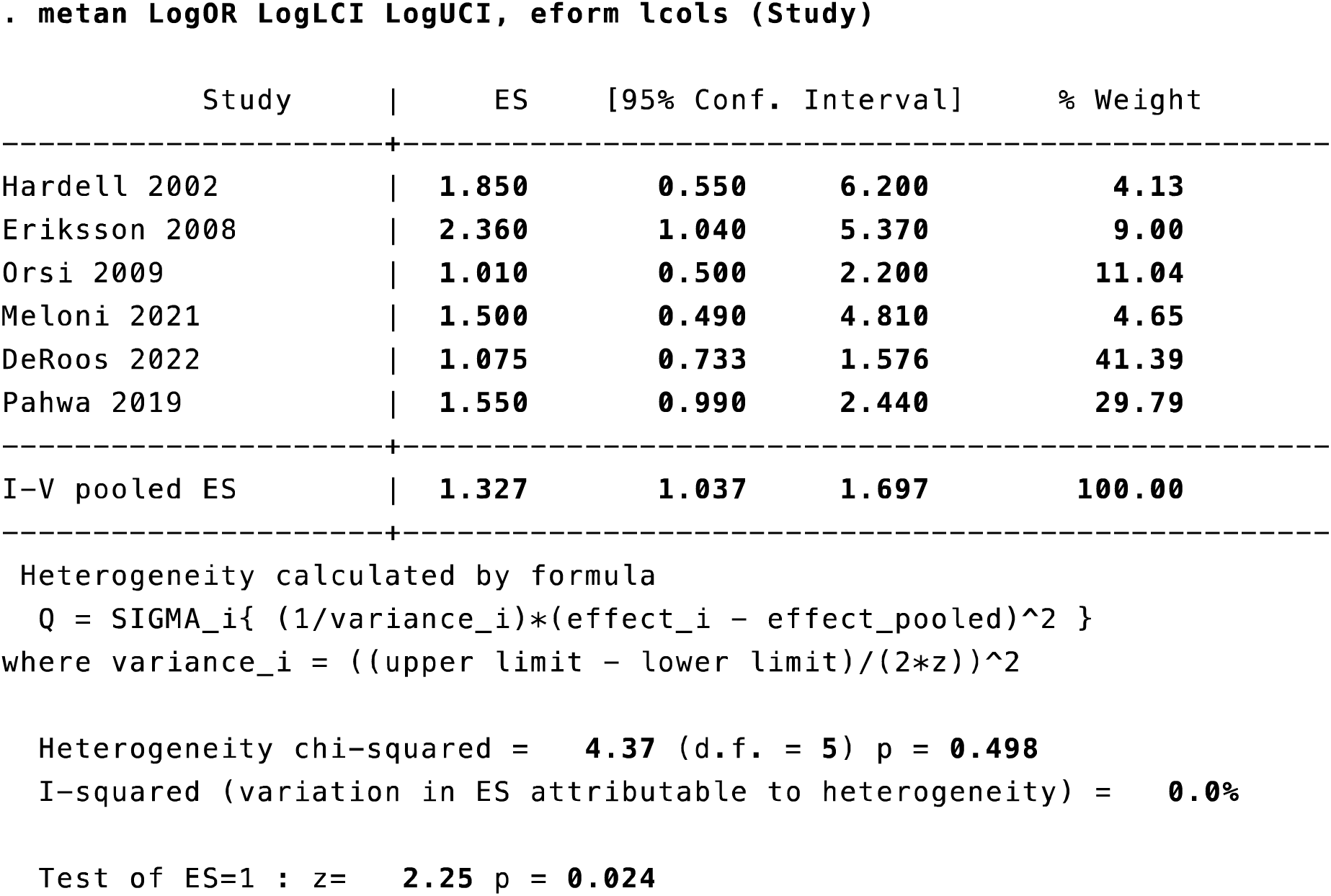
Fixed Effects Meta-Analysis Results Table for Highest Exposure Category with Cumulative Exposure from Meloni and Pahwa removing Leon and Poh

**Figure 10:**
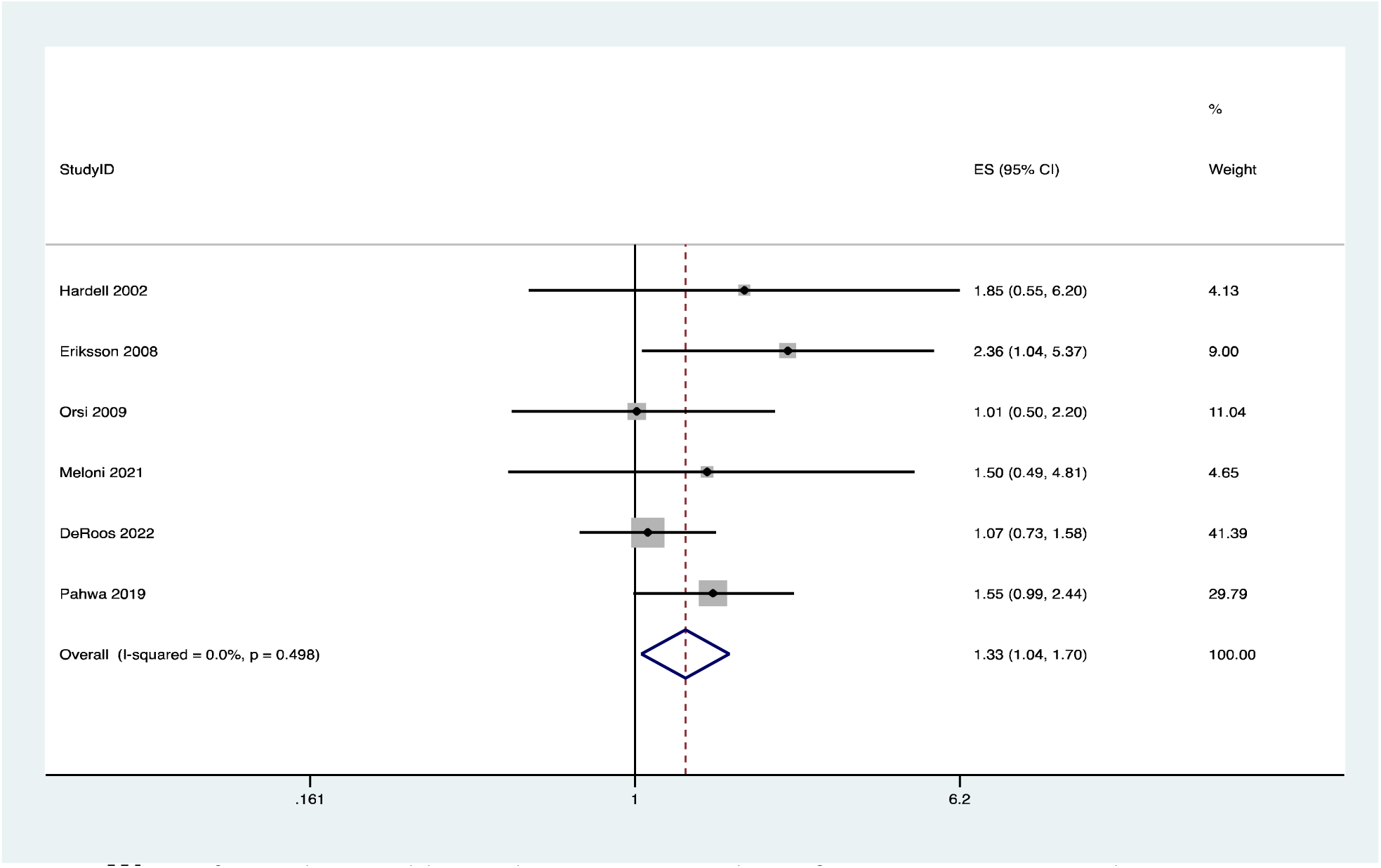
Forest Plot Fixed Effects Meta-Analysis for highest exposure category with Cumulative Exposure from Meloni and Pahwa removing Leon and Poh

We performed two additional sensitivity analyses for ever-exposure and maximum exposure to glyphosate by removing Hardell study [7] since this study included hairy cell leukemia (HCL), not always considered a form of NHL, and found similar results for all estimates (not presented here).

##### Investigation of Publication Bias

The standard funnel plot for the main analysis for the relationship of ever exposure to NHL is shown in figure 13, with the Egger regression line. On visual inspection and using the Egger test for funnel plot asymmetry it appears that there was no publication bias, or other small/imprecise study effect, (Egger test p=0.423) indicating no publication bias. Note, the Egger test and funnel plot is underpowered, and potentially misleading when you have less than 10 studies. Therefore, this statistical test should be viewed with caution.

**Figure 13:**
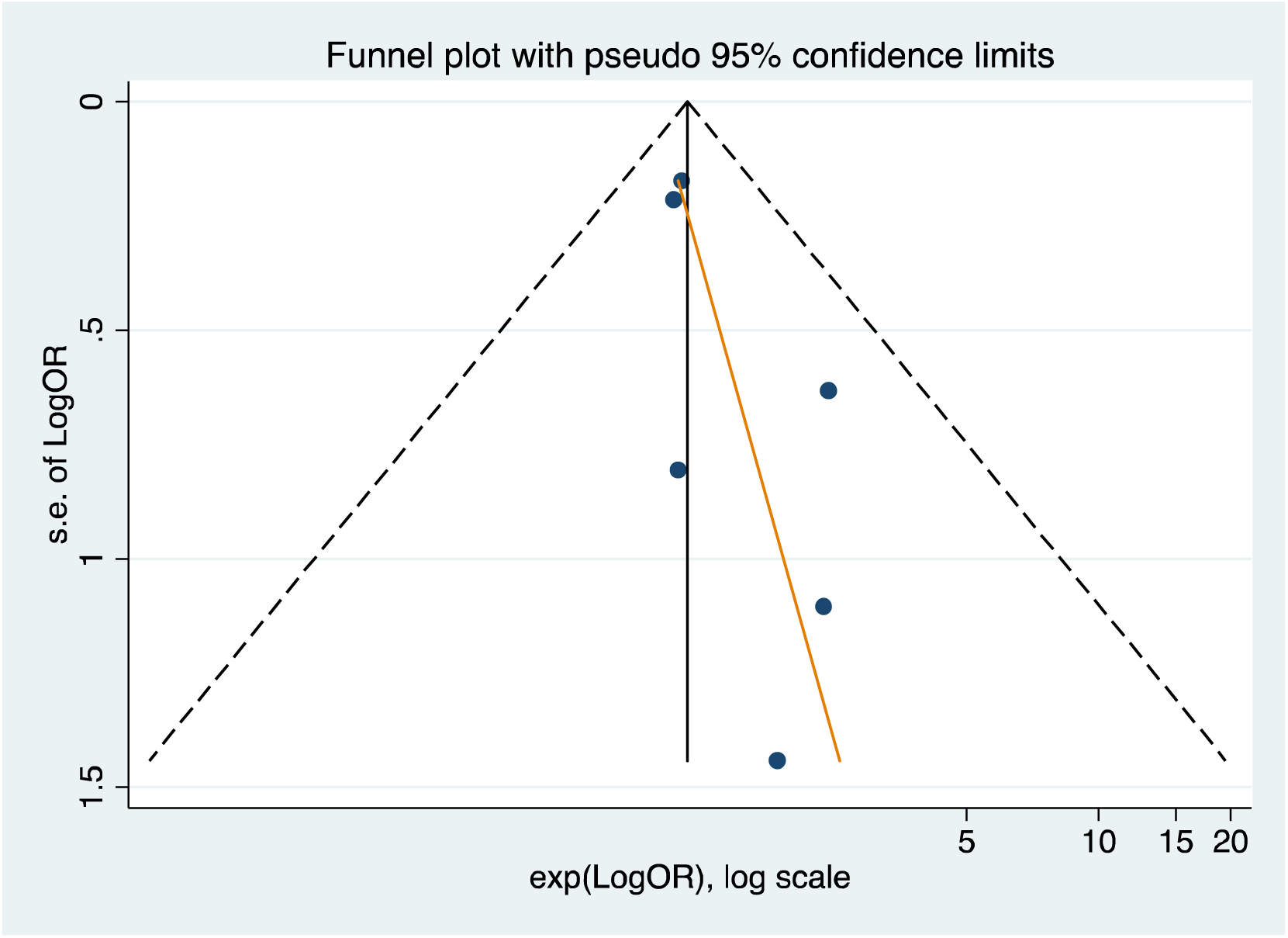
Standard funnel plot for ever exposure data for all non-duplicate included studies.

To confirm the role of p-values, true publication bias, we also ran contour-enhanced funnel lots, which can determine if lack of publication of imprecise studies due to p-value cutoffs (see figure 14). This funnel plot shows that it is possible that imprecise negative studies are missing to the left of the lower bunch of studies on the plot, but it also appears that the more precise (larger) studies show some negative results (which in this case include the pooled studies of several smaller studies), making the risk of true publication bias due to missing small studies unlikely.

**Figure 14:**
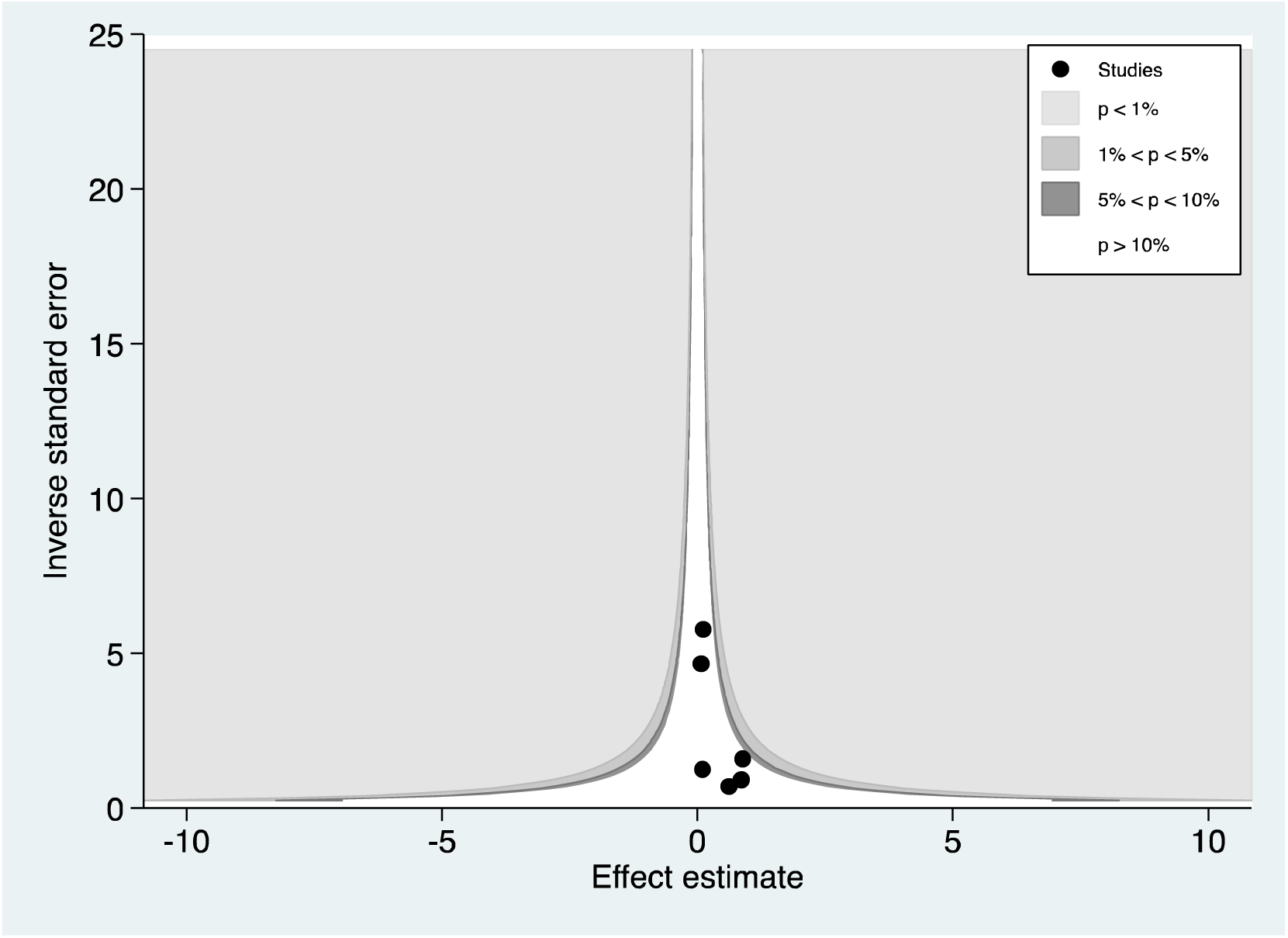
Contour-enhanced plot for ever exposure data for all non-duplicate included studies.

Therefore, it is possible that the small study effects I see in the standard funnel plot are due to methodologic strengths in these imprecise (smaller individual) studies. Overall, this analysis is inconclusive for the presence of true publication bias due to the small number of studies included.

Next, we performed the same funnel plots for the highest level of exposure, using the cumulative exposure values where available. Figure 15 shows the standard funnel plot with the Egger regression line, that was non-statistically significant for funnel plot asymmetry (p=0.108).

**Figure 15:**
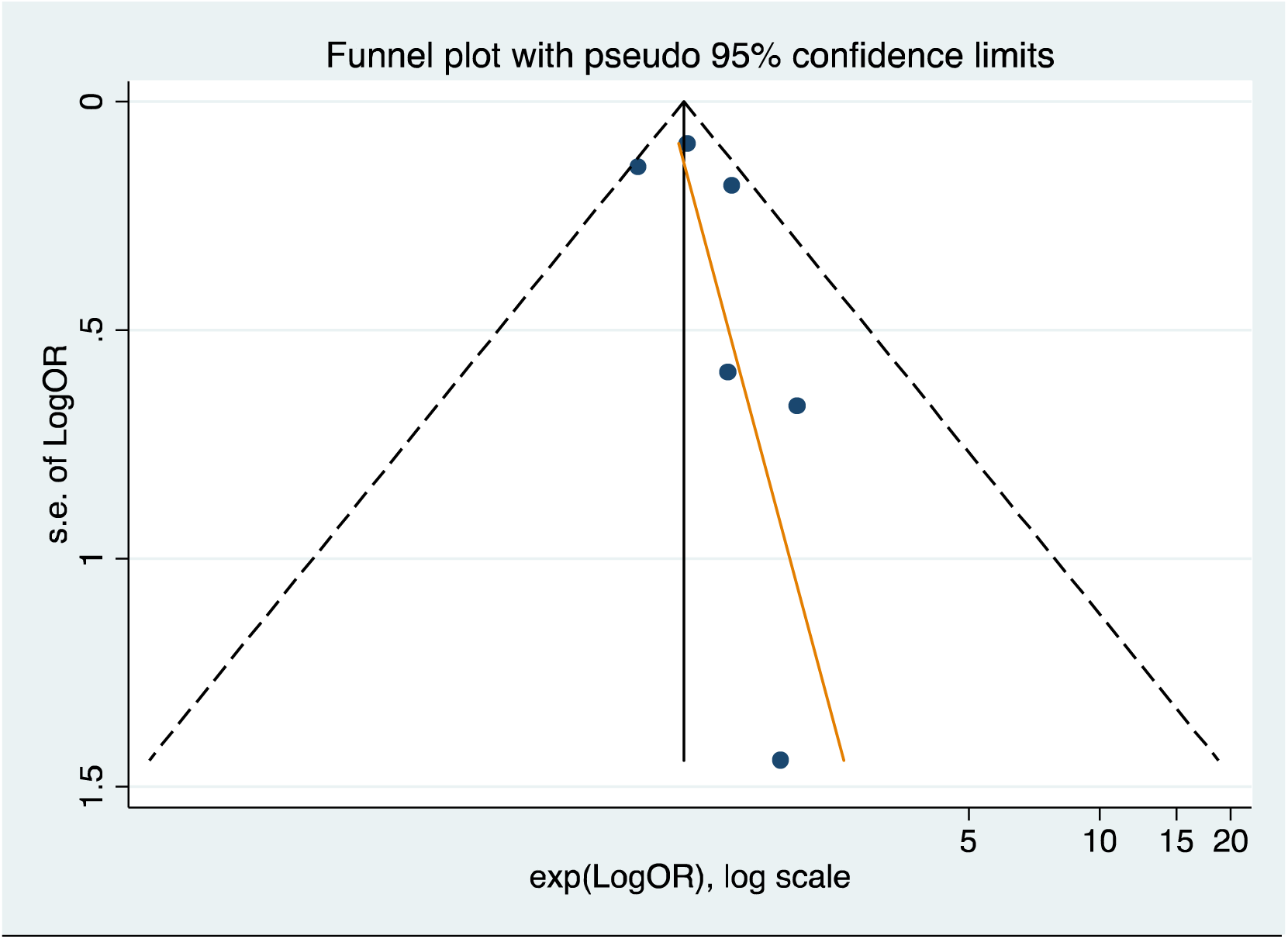
Standard funnel plot for highest level of exposure data for all non-duplicate included studies.

The contour-enhanced funnel plot is shown in figure 16, which has a very different interpretation as for the standard funnel plot, in this case the risk of true publication bias appears to even lower and instead the asymmetry is likely due to robust small study effect, true effects and not true publication bias. Though, for both sets of funnel plots the power is still low so the interpretation of these plots should be made with caution.

**Figure 16:**
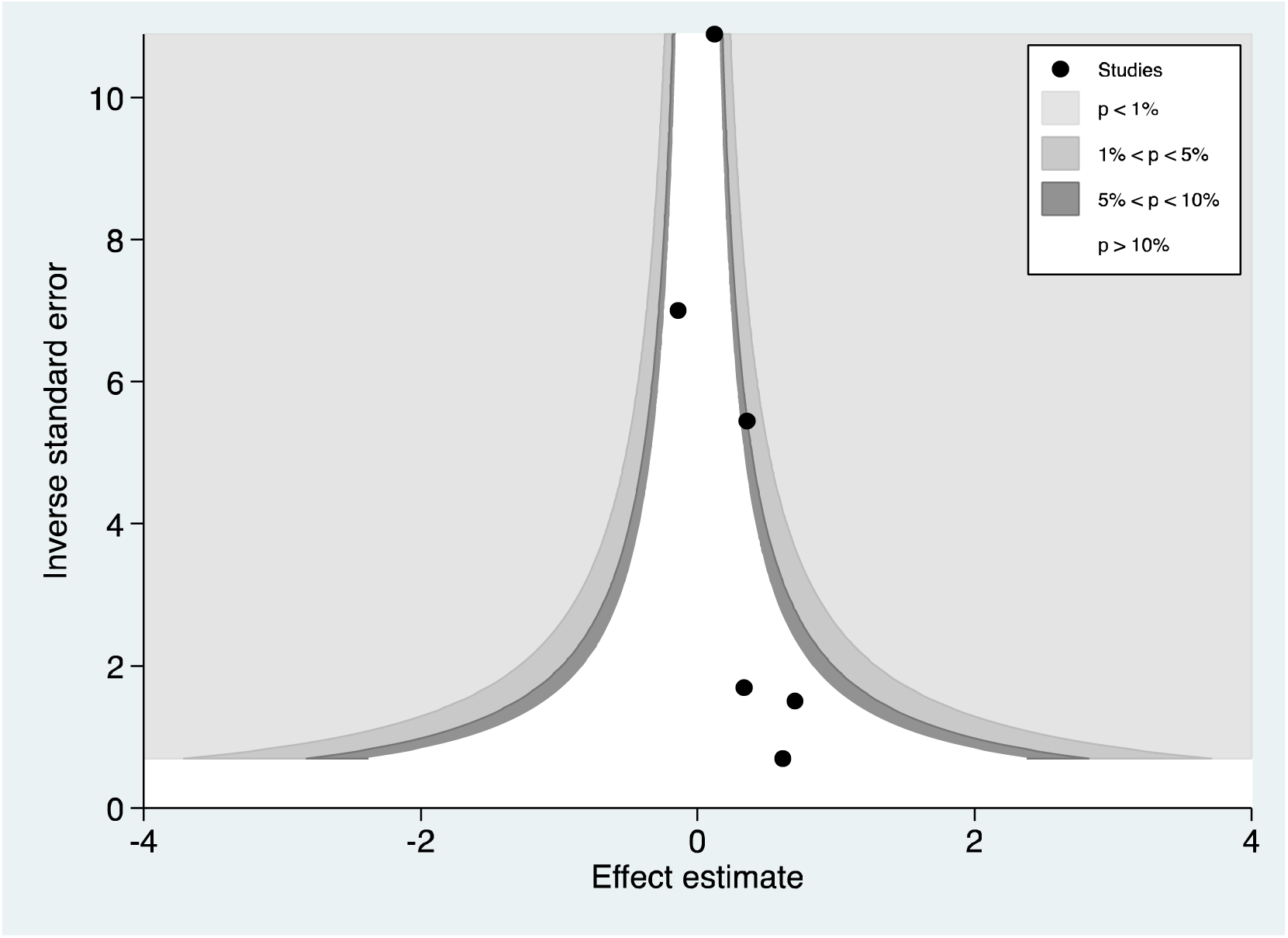
Contour-enhanced funnel plot for highest level of exposure data for all non-duplicate included studies.

##### Meta-Regression

We then performed meta-regression analyses for the influence of study quality on the estimated of treatment effect of the two main analyses including all non-overlapping studies, for ever-exposed and for the highest level of exposure, excluding Poh and Leon due to their use of hazards ratios. Results are shown in figures 17 and 18.

**Figure 17:**
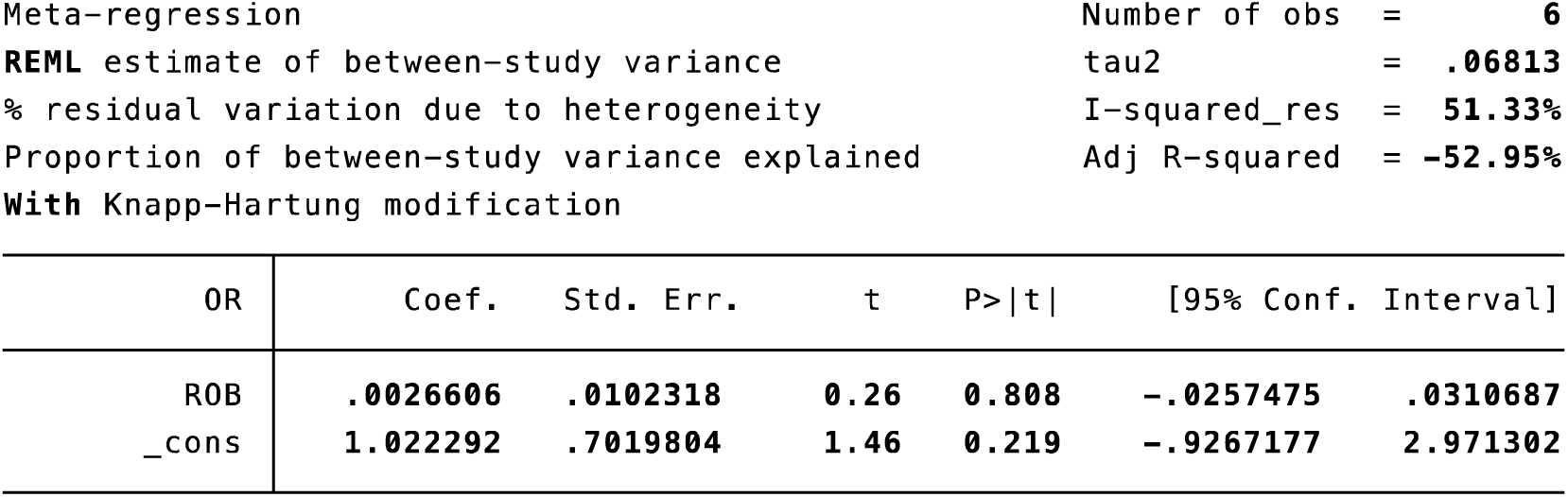
Meta-regression exploring variation in ever-exposed meta-analytic estimate by risk of bias

**Figure 18:**
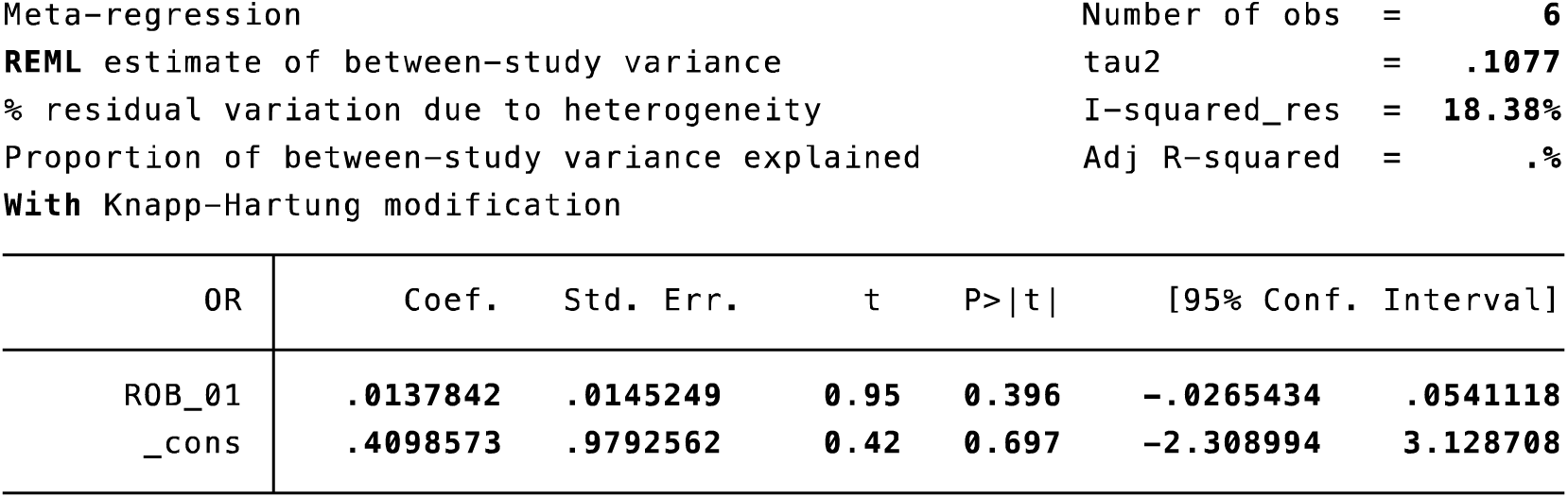
Meta-regression exploring variation in highest exposure meta-analytic estimate by risk of bias

The meta-regression analyses both revealed that the meta-analytic treatment effect estimates for the relationship of glyphosate exposure with NHL did not change across varying levels of risk of bias of the included studies. Furthermore, we performed several additional meta-regression analyses grouping low and moderate risk of bias and comparing this to high risk of bias studies, and again, for low risk of comparing this to moderate combined with high risk of bias finding no significant effects. Therefore, the meta-analytic effect estimates presented above are robust to varying risk of bias of the included studies.

##### Grade Assessment

The GRADE table is shown in table 4. The two assessors agreed on the GRADE assessment. Across all analyses, the overall evidence, using the grade approach, is moderate.

Systematic reviews of observational studies start as low evidence and can go down or up. In this case I did not rate the evidence down for any of the certainty assessments described by the GRADE working group [33] and instead increased the GRADE of evidence to moderate due to the dose-response relationship shown across the included studies and as evidenced by the increased ORs when going from ever-exposure to the highest level of exposure in days to cumulative exposure metrics. Therefore, the overall the GRADE of evidence is moderate. In the GRADE approach high GRADE evidence is typically limited to systematic reviews and meta-analyses of randomized trials only.

## Discussion

The results from our main analyses suggest an increased risk of 18% to 24%, from the analyses without Leon and Poh, the most valid analyses (least risk of type 1 errors), for NHL in those ever exposed to Glyphosate at any point in their life. The results from our main analyses also suggest an increased risk of 33% to 47% for NHL, again from the analyses without Leon and Poh, in those exposed to Glyphosate at the various highest levels of exposure definitions used in the individual studies. These findings were robust in the face of sensitivity analyses, do not appear to be influenced by publication bias and do not vary by study quality. Overall, there is a moderate GRADE of evidence that exposure to glyphosate increased the odds of NHL in humans in those ever exposed or most highly exposed across a very large sample of patients, in various study designs, geographic regions and by varying research teams.

Compared with prior systematic reviews [3,11,13,19,20,27], the present investigation provides a methodologically rigorous synthesis of the evidence on glyphosate exposure and NHL. Earlier reviews varied substantially in scope, eligibility criteria, exposure definitions, and analytic strategies, which likely contributed to the inconsistent conclusions in this literature.

Some prior reviews reported modest positive associations for ever-use of glyphosate or stronger associations for higher or more prolonged exposure, whereas others concluded that the overall evidence was weak, inconsistent, or compatible with no clear association. In particular, reviews emphasizing highest cumulative exposure tended to report more pronounced positive associations, while reviews focused on ever-exposure estimates generally found smaller effects or null results [3,11,13,19,20,27]. Our findings help reconcile these differences.

A key distinction of our systematic review is that it addressed several important limitations of earlier reviews [3,11,13,19,20,27]. First, we incorporated a broader and more current body of literature and took explicit steps to avoid double-counting overlapping study populations in the quantitative synthesis. Second, we applied formal risk-of-bias assessment and certainty-of-evidence evaluation, which were limited or absent in most previous reviews. Third, we explored the impact of varying risk of bias across the included studies, showing little impact. Fourth, we conducted a series of sensitivity analyses to examine the robustness of findings to alternative exposure definitions, inclusion of hazard-ratio estimates, overlapping cohorts, and potentially influential analytic decisions. These steps provide a clearer basis for interpreting why past meta-analyses reached differing conclusions and increase the robustness of our review findings.

Here we have completed the most up to date, largest and rigorous systematic review and meta-analysis ever completed on the question, “Does exposure to glyphosate increase the risk or not for NHL?”. Overall, most meta-analyses and sensitivity analyses performed yield the same answer, which is yes. The evidence is very good, and consistent across 20 separate data sets published in 9 studies, that glyphosate increases the risk of NHL in those ever exposed and even a greater risk in those with the highest measured exposures. Also, while some of the included studies had methodologic flaws, the risk of bias of the included studies did not have any influence on these findings, whether treated continuously or categorically. The risk of publication bias is not likely for the reasons described above, and in fact the small study effects seen may have been due to reasons other than true publication bias. As noted, the power of the test for publication bias is low due to the small number of included studies used (n=6). Overall, we have high confidence in the results presented above. Future research on this question could explore the varying risk for subtypes of NHL and the role of additional effect modifiers in carefully powered analyses.

The scientific evidence available from published human subjects epidemiologic research points to an increased risk of NHL in those ever exposed to and for high levels of exposure to glyphosate. We have high confidence that any follow-up research on this question will yield similar findings.

## Clinical Trial Number

Not applicable.

## Data Availability

All data produced in the present study are available upon reasonable request to the authors

